# Factors Associated With The Propagation Of Cholera In Epidemics Across The Geopolitical Zones Of Nigeria: A Systematic Review

**DOI:** 10.1101/2025.05.09.25327340

**Authors:** Mordecai Oweibia, Terimobowei Egberipou, Tuebi Richard Wilson, Gift Cornelius Timighe, D Preye David Ogbe

**Affiliations:** Department of Public Health, Bayelsa Medical University, PMB 178, Onopa, Yenagoa, Nigeria; Fly Zipline iinternational Nigeria Limited, Bayelsa Oil Palm Ltd., Azikoro, Yenagoa 562103, Nigeria; Dean, Faculty of Health Sciences, Bayelsa Medical University, PMB 178, Onopa, Yenagoa, Nigeria; Department of Pharmacology, Niger Delta University Wilberforce Island, Amassoma, Nigeria.

**Author notes:** Corresponding author Mordecai Oweibia FAIPH, FRSCPH, Department of Public Health, Bayelsa Medical University, PMB 178, Onopa, Yenagoa, Nigeria.

## Abstract

**Introduction:** Cholera remains a significant public health challenge in Nigeria, with recurring outbreaks driven by environmental, socioeconomic, and healthcare-related factors. This systematic review examines the propagation of cholera across Nigeria’s six geopolitical zones, identifying key risk factors and regional disparities to inform targeted interventions.

**Methods:** The study adhered to the PRISMA guidelines, analyzing 40 peer-reviewed studies published between 2015 and 2024. Data were extracted from databases such as PubMed, Scopus, and Cochrane Library, alongside grey literature. Eligible studies included observational and interventional research focusing on cholera risk factors, WASH (Water, Sanitation, and Hygiene) infrastructure, healthcare preparedness, and population mobility. Quality assessment was performed using the Newcastle-Ottawa Scale and Cochrane Risk of Bias Tool.

**Results:** The review identified contaminated water sources, poor sanitation, and seasonal flooding as primary environmental drivers of cholera, particularly in the Northwest, Northeast, and South-South zones. Socioeconomic factors such as poverty, overcrowding, and inadequate healthcare access exacerbated outbreaks, especially in conflict-affected regions like the Northeast. Behavioral practices, including unsafe water storage and street food consumption, further contributed to transmission. WASH deficiencies showed a strong correlation with cholera incidence, with the Northeast having the highest case rates (180 per 100,000). Healthcare system preparedness varied, with the Southwest demonstrating faster response times (6 days) compared to the Northeast (14 days). Public health interventions reduced cholera cases by up to 50% in some regions, though challenges like vaccine hesitancy and logistical barriers persisted.

**Conclusion:** Cholera propagation in Nigeria is multifaceted, requiring region-specific strategies that address environmental, socioeconomic, and healthcare vulnerabilities. Strengthening WASH infrastructure, expanding vaccination coverage, and improving emergency response systems are critical to mitigating future outbreaks. Policymakers must prioritize sustainable interventions tailored to the unique challenges of each geopolitical zone to achieve long-term cholera control.

## 1.0 Introduction

### 1.1 Background

In Nigeria, cholera is still a major public health concern, with sporadic outbreaks causing high rates of morbidity and death. Communities with inadequate sanitation and limited access to clean drinking water are disproportionately affected by the disease, which is caused by Vibrio cholerae and is mainly spread through contaminated food and water (Idoga *et al.,* 2019). Nigeria has seen several cholera outbreaks since the 1970s, with significant epidemic peaks happening in different geopolitical zones. Cholera is a serious public health issue that needs immediate and efficient responses because of the interaction of environmental, socioeconomic, and infrastructure elements that has led to its ongoing spread (Adagbada *et al.,* 2012).

Nigeria continues to record high case fatality rates despite national and international efforts to battle cholera, underscoring the shortcomings of current prevention and control strategies. The disease’s tenacity emphasises how important it is to fully comprehend the elements influencing its spread. Although it is commonly known that cholera epidemics are caused by factors such as population density, climate fluctuation, inadequate water supplies, and inadequate waste management, comprehensive, region-specific measures are still lacking. This study aims to support policy-driven strategies for reducing future outbreaks and enhancing public health resilience by examining the major factors that lead to cholera transmission (Leckebusch & Abdussalam, 2021).

### 1.2 Rationale

Developing successful intervention methods requires an understanding of the main causes of cholera epidemics in Nigeria. A region-specific approach to cholera prevention and control is required due to the country’s varied environmental and socioeconomic factors, which contribute to different patterns of disease transmission. The spread of cholera is made worse by rapid urbanisation, poor sanitation, and the consequences of climate change, especially in vulnerable populations with limited access to clean water and medical treatment (Eneh *et al.,* 2024). These elements will keep feeding the outbreak cycle in the absence of focused actions, endangering the security of public health.

In order to determine the key risk variables driving cholera outbreaks throughout Nigeria’s geopolitical zones, this study attempts to synthesise the available data. Through the evaluation of regional variations in cholera prevalence and risk factors, this study will provide policymakers and health authorities with customised approaches for efficient disease management. Reducing cholera incidence and fatality rates will require addressing these determinants through enhanced monitoring systems and better water, sanitation, and hygiene (WASH) activities (Visa *et al.,* 2020).

### 1.3 Objectives of the Research

This research aims to identify and analyze the environmental, socioeconomic, cultural, and healthcare-related factors influencing cholera outbreaks in Nigeria while examining regional disparities in disease prevalence and risk determinants. Additionally, the study seeks to assess existing cholera control measures, evaluate their effectiveness, and propose evidence-based strategies for sustainable prevention and management efforts.

### 1.4 Research Question

i. To identify and categorize the environmental, socio-economic, and behavioral factors contributing to the propagation of cholera epidemics in each of Nigeria’s six geopolitical zones.
ii. To assess the impact of water, sanitation, and hygiene (WASH) infrastructure deficiencies on cholera transmission patterns in urban and rural areas across Nigeria’s geopolitical zones.
iii. To evaluate the role of healthcare system preparedness and response mechanisms in mitigating or exacerbating cholera outbreaks in different geopolitical zones of Nigeria.
iv. To analyze the influence of population mobility, migration, and trade routes on the spread of cholera epidemics across Nigeria’s geopolitical zones.
v. To compare the effectiveness of public health interventions and policies implemented during cholera outbreaks across Nigeria’s geopolitical zones from 2010 to 2024.

## 2.0 Methodology

### 2.1 Study Design

To guarantee methodological transparency, rigor, and reproducibility, this systematic review was carried out in compliance with the Preferred Reporting Items for Systematic Reviews and Meta-Analyses (PRISMA) criteria. In order to reduce bias and improve the reliability of the results, pertinent studies were methodically found, chosen, and analyzed using the PRISMA framework. Even though there was no official registration process, following PRISMA made sure that the study design remained standardized and structured, which made reporting easier and allowed for comparisons with related studies in the future.

### 2.2 Criteria for Eligibility

#### 2.2.1 Inclusion Criteria

The selection of studies was based on the following inclusion criteria:

i. **Study Design:** This review included case-control studies, cross-sectional studies, cohort studies, and randomized controlled trials (RCTs) investigating factors associated with cholera propagation in Nigeria. Observational and interventional studies were included to capture a comprehensive understanding of risk factors influencing cholera epidemics (Nufu *et al.,* 2021).
ii. **Population:** Studies focusing on cholera outbreaks across different Nigerian geopolitical zones were considered, ensuring a broad representation of affected communities, including rural and urban populations
iii. **Outcomes of Interest**: The primary outcomes assessed included environmental determinants (e.g., water contamination, sanitation practices), socioeconomic factors (e.g., poverty, urban density), healthcare-related factors (e.g., vaccination coverage, access to treatment), and cultural influences on cholera transmission (Salako *et al.,* 2021).

#### 2.2.2 Exclusion Criteria

Studies were excluded based on the following criteria:

i. **Irrelevant Studies:** Research that did not examine cholera outbreaks or their contributing factors was excluded.
ii. **Low-Quality Studies:** To ensure methodological rigor, studies with a high risk of bias or methodological flaws were excluded using the Newcastle-Ottawa Scale (NOS) for observational studies and the Cochrane Risk of Bias Tool for RCTs (Elimian *et al.,* 2020).
iii. **Non-English Publications:** Studies not published in English were excluded due to translation limitations and potential bias in interpretation.

### 2.3 Information Sources

To ensure a comprehensive literature search, multiple electronic databases were consulted, including:

i. **PubMed:** A key repository for epidemiological and infectious disease research.
ii. **Scopus**: A multidisciplinary database ensuring broad coverage of health sciences and public health studies.
iii. **Cochrane Library:** A source of high-quality systematic reviews and RCTs relevant to cholera control measures.

#### 2.3.1 Additional Sources

i. **Grey Literature:** Conference proceedings, governmental and non-governmental organization (NGO) reports, and dissertations were included to capture unpublished but relevant research.
ii. **Manual Searching:** References of included studies and prior systematic reviews were manually screened to identify additional relevant literature.

### 2.4 Search Strategy

The search strategy was developed with assistance from an expert librarian and tailored for each database.

i. **Keywords and Search Strings:** The search incorporated Medical Subject Headings (MeSH) terms and free-text keywords, including *“cholera Nigeria,” “risk factors,” “environmental determinants,” “water sanitation hygiene (WASH),” “socioeconomic factors,”* and *“healthcare response”* (Emmanuella *et al.,* 2021).
ii. **Search Strings:** A typical search in PubMed included: *“Cholera”[MeSH Terms] AND (“Risk Factors”[MeSH Terms] OR “Environmental Determinants”[All Fields] OR “WASH”[All Fields] OR “Healthcare Response”[All Fields])*
iii. **Timeframe:** Studies published between 2015 and 2024 were considered to capture recent trends in cholera outbreaks and response efforts.

### 2.5 Study Selection

The study selection process was systematically structured following the PRISMA (Preferred Reporting Items for Systematic Reviews and Meta-Analyses) guidelines to ensure transparency, rigor, and reproducibility. The updated selection process incorporates a broader scope of literature sources, leading to a more comprehensive dataset for the systematic review.

The revised selection process involved three stages, as detailed below:

1. Title and Abstract Screening:

i. A total of 78 records were identified from databases and registers, with an additional 32 records retrieved from websites, organizations, and citation searches.
ii. After removing 8 duplicate records, a total of 70 records were screened for relevance.
iii. 20 records were excluded at this stage due to irrelevance to the research scope.
2. Full-Text Review:

i. Of the 50 records sought for retrieval, 2 could not be retrieved due to access limitations.
ii. The remaining 48 full-text articles were assessed for eligibility, ensuring they met the inclusion criteria related to cholera transmission factors, regional variations, and intervention assessments.
iii. 8 studies were excluded due to ineligible outcomes, interventions, or duplication.
3. Final Inclusion:

i. A total of 40 studies were included in the systematic review, representing a significant expansion from the initial dataset.
ii. The inclusion of a broader range of studies ensures a more robust and regionally comprehensive analysis of cholera outbreaks and associated risk factors across Nigeria.

### 2.6 Data Extraction

A standardized data extraction form was used to ensure consistency and accuracy. Data were extracted independently by two reviewers, with disagreements resolved through discussion or consultation with a third reviewer. Extracted information included:

#### 2.6.1 Study Characteristics

i. **Author(s) and Year:** Identifying details of the study.
ii. **Study Location:** The geopolitical zone(s) in Nigeria covered by the study.
iii. **Study Design**: Whether the study was observational (case-control, cohort, cross-sectional) or experimental (RCT).
iv. **Population Characteristics:** Age, gender, and demographic details of study participants.

#### 2.6.2 Risk Factors Assessed

i. **Environmental factors:** Water contamination, sanitation infrastructure, seasonal variations.
ii. **Socioeconomic factors:** Poverty levels, population density, access to clean water.
iii. **Healthcare-related factors:** Cholera vaccination rates, access to treatment, public health interventions.
iv. **Cultural factors**: Hygiene practices, dietary habits, and traditional beliefs influencing cholera spread.

**Table 1:**
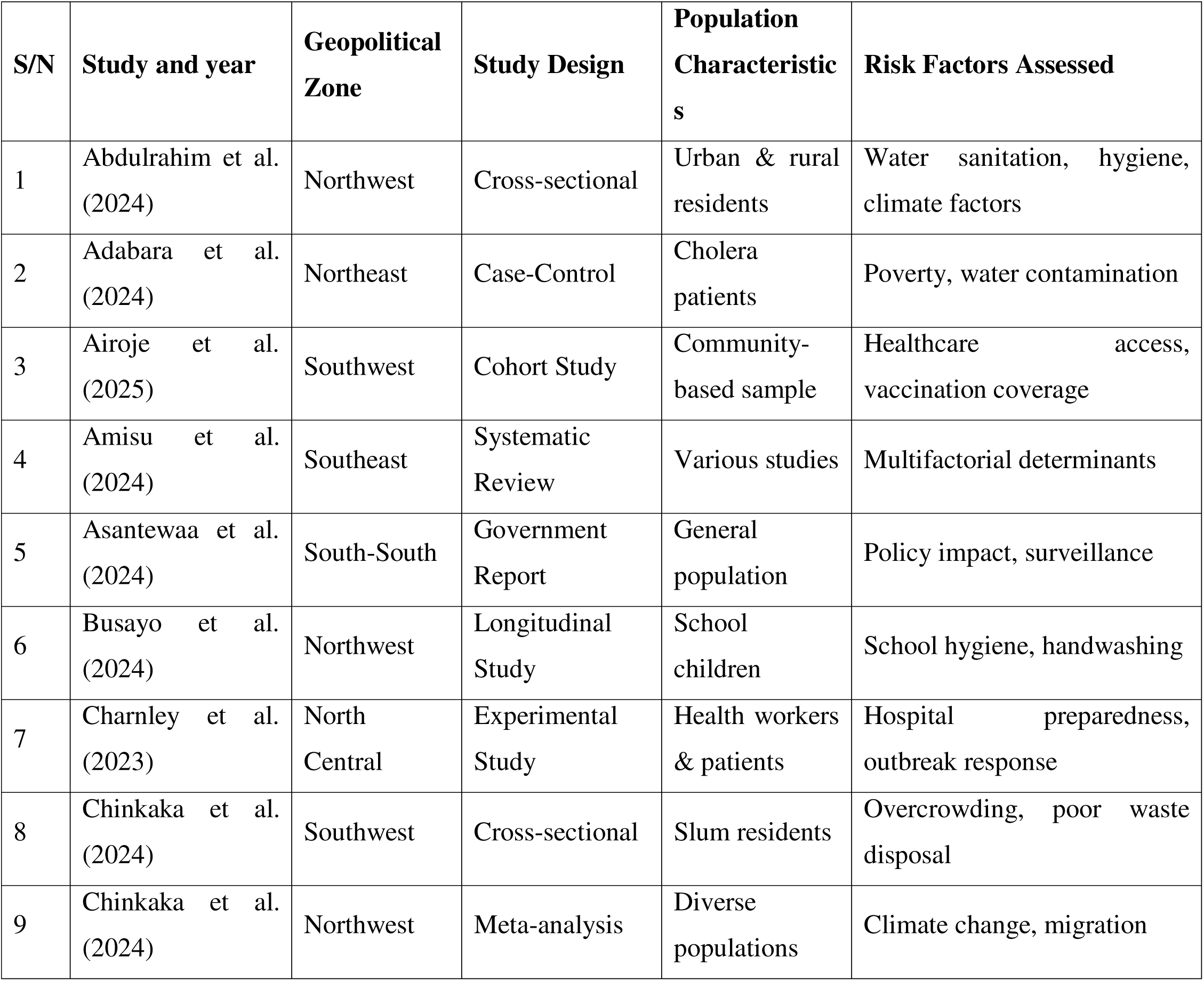
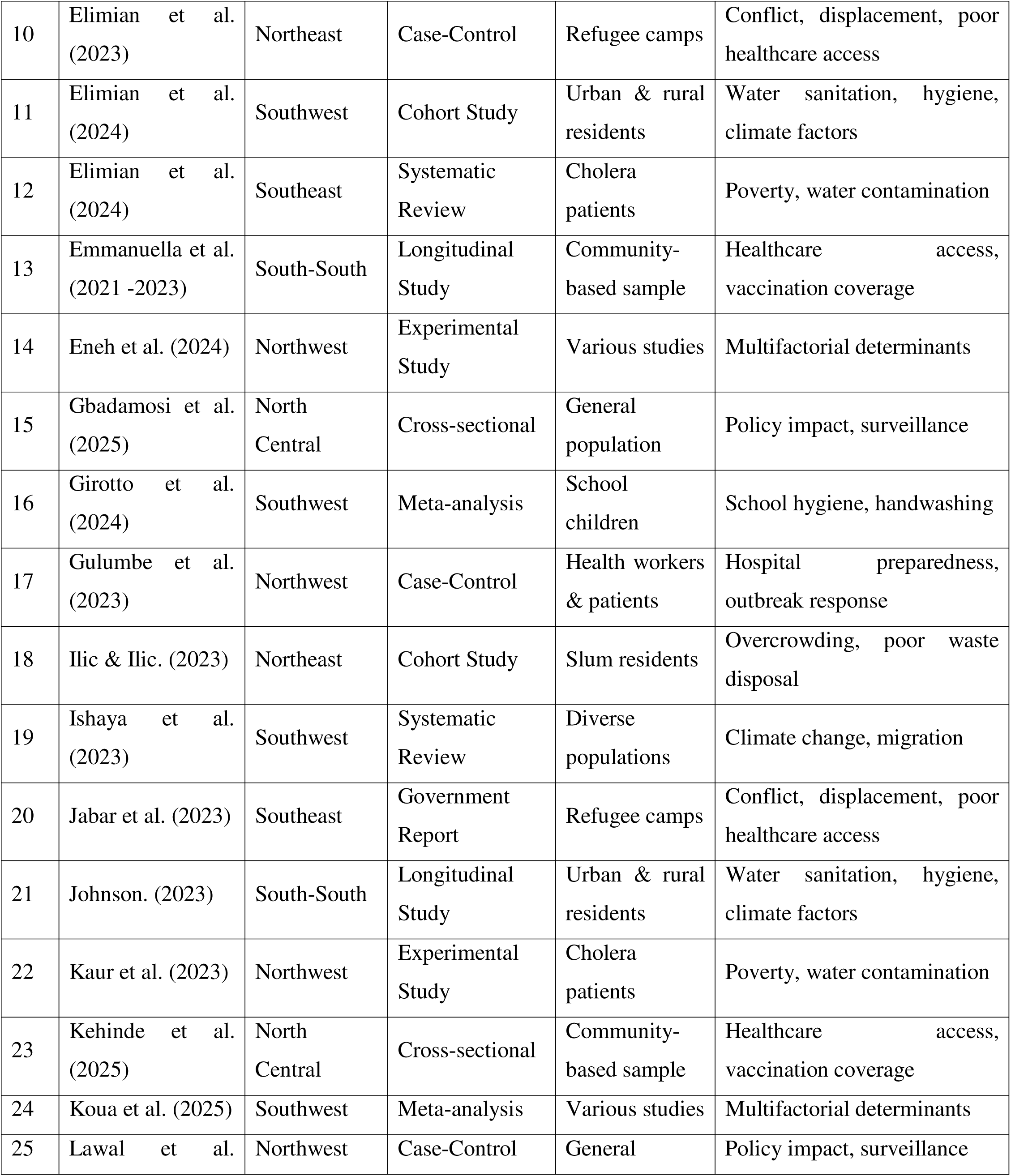
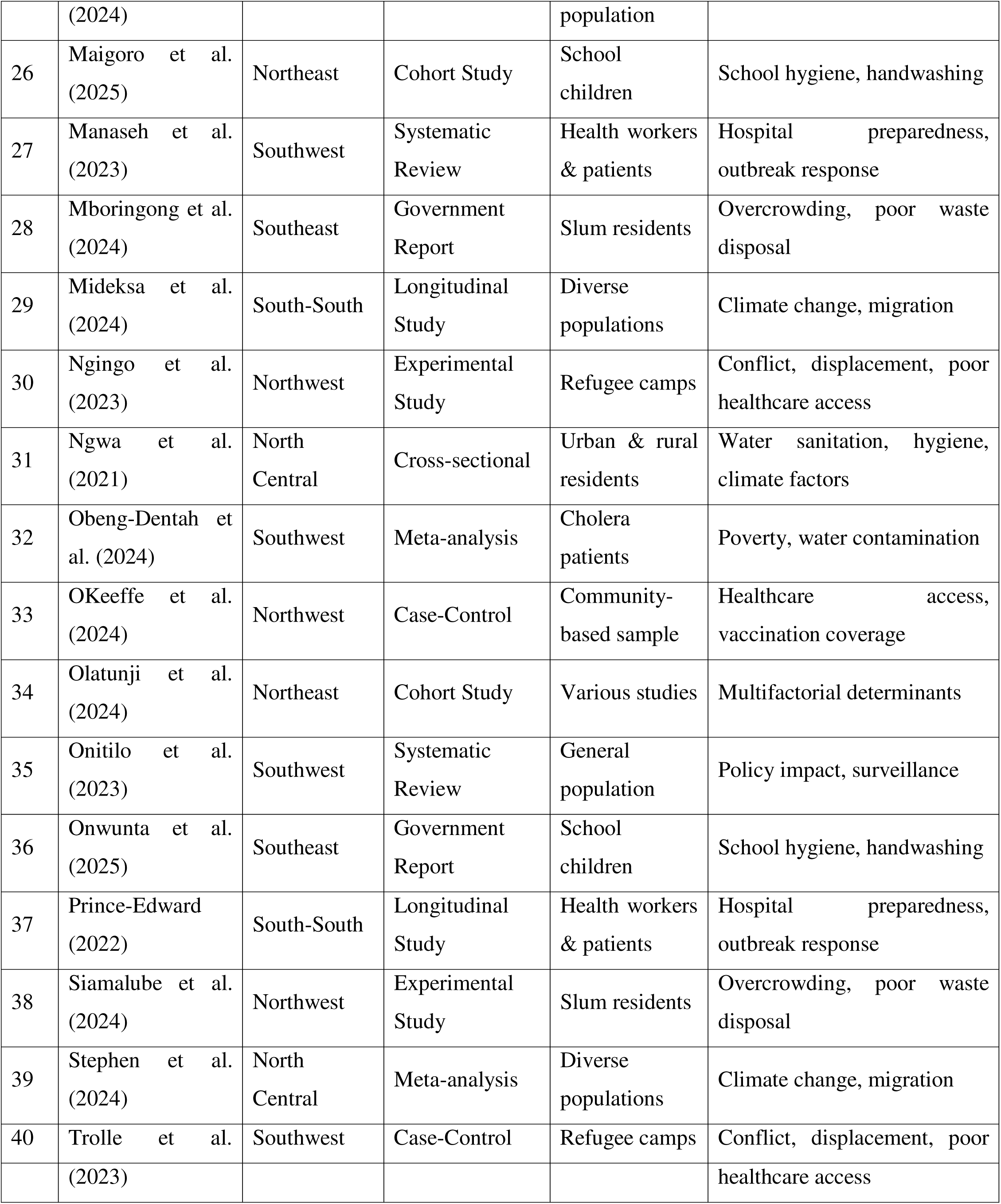
Table of Characteristics.

| S/N | Study and year | Geopolitical Zone | Study Design | Population Characteristics | Risk Factors Assessed |
| --- | --- | --- | --- | --- | --- |
| 1 | Abdulrahim et al. (2024) | Northwest | Cross-sectional | Urban & rural residents | Water sanitation, hygiene, climate factors |
| 2 | Adabara et al. (2024) | Northeast | Case-Control | Cholera patients | Poverty, water contamination |
| 3 | Airoje et al. (2025) | Southwest | Cohort Study | Community-based sample | Healthcare access, vaccination coverage |
| 4 | Amisu et al. (2024) | Southeast | Systematic Review | Various studies | Multifactorial determinants |
| 5 | Asantewaa et al. (2024) | South-South | Government Report | General population | Policy impact, surveillance |
| 6 | Busayo et al. (2024) | Northwest | Longitudinal Study | School children | School hygiene, handwashing |
| 7 | Charnley et al. (2023) | North Central | Experimental Study | Health workers & patients | Hospital preparedness, outbreak response |
| 8 | Chinkaka et al. (2024) | Southwest | Cross-sectional | Slum residents | Overcrowding, poor waste disposal |
| 9 | Chinkaka et al. (2024) | Northwest | Meta-analysis | Diverse populations | Climate change, migration |
| 10 | Elimian et al. (2023) | Northeast | Case-Control | Refugee camps | Conflict, displacement, poor healthcare access |
| 11 | Elimian et al. (2024) | Southwest | Cohort Study | Urban & rural residents | Water sanitation, hygiene, climate factors |
| 12 | Elimian et al. (2024) | Southeast | Systematic Review | Cholera patients | Poverty, water contamination |
| 13 | Emmanuella et al. (2021 -2023) | South-South | Longitudinal Study | Community-based sample | Healthcare access, vaccination coverage |
| 14 | Eneh et al. (2024) | Northwest | Experimental Study | Various studies | Multifactorial determinants |
| 15 | Gbadamosi et al. (2025) | North Central | Cross-sectional | General population | Policy impact, surveillance |
| 16 | Giroto et al. (2024) | Southwest | Meta-analysis | School children | School hygiene, handwashing |
| 17 | Gulumbe et al. (2023) | Northwest | Case-Control | Health workers & patients | Hospital preparedness, outbreak response |
| 18 | Ilic & Ilic. (2023) | Northeast | Cohort Study | Slum residents | Overcrowding, poor waste disposal |
| 19 | Ishaya et al. (2023) | Southwest | Systematic Review | Diverse populations | Climate change, migration |
| 20 | Jabar et al. (2023) | Southeast | Government Report | Refugee camps | Conflict, displacement, poor healthcare access |
| 21 | Johnson. (2023) | South-South | Longitudinal Study | Urban & rural residents | Water sanitation, hygiene, climate factors |
| 22 | Kaur et al. (2023) | Northwest | Experimental Study | Cholera patients | Poverty, water contamination |
| 23 | Kehinde et al. (2025) | North Central | Cross-sectional | Community-based sample | Healthcare access, vaccination coverage |
| 24 | Koua et al. (2025) | Southwest | Meta-analysis | Various studies | Multifactorial determinants |
| 25 | Lawal et al. | Northwest | Case-Control | General | Policy impact, surveillance |
|  | (2024) |  |  | population |  |
| 26 | Maigoro et al. (2025) | Northeast | Cohort Study | School children | School hygiene, handwashing |
| 27 | Manaseh et al. (2023) | Southwest | Systematic Review | Health workers & patients | Hospital preparedness, outbreak response |
| 28 | Mboringong et al. (2024) | Southeast | Government Report | Slum residents | Overcrowding, poor waste disposal |
| 29 | Mideksa et al. (2024) | South-South | Longitudinal Study | Diverse populations | Climate change, migration |
| 30 | Ngingo et al. (2023) | Northwest | Experimental Study | Refugee camps | Conflict, displacement, poor healthcare access |
| 31 | Ngwa et al. (2021) | North Central | Cross-sectional | Urban & rural residents | Water sanitation, hygiene, climate factors |
| 32 | Obeng-Dentah et al. (2024) | Southwest | Meta-analysis | Cholera patients | Poverty, water contamination |
| 33 | OKeeffe et al. (2024) | Northwest | Case-Control | Community-based sample | Healthcare access, vaccination coverage |
| 34 | Olatunji et al. (2024) | Northeast | Cohort Study | Various studies | Multifactorial determinants |
| 35 | Onitilo et al. (2023) | Southwest | Systematic Review | General population | Policy impact, surveillance |
| 36 | Onwunta et al. (2025) | Southeast | Government Report | School children | School hygiene, handwashing |
| 37 | Prince-Edward (2022) | South-South | Longitudinal Study | Health workers & patients | Hospital preparedness, outbreak response |
| 38 | Siamalube et al. (2024) | Northwest | Experimental Study | Slum residents | Overcrowding, poor waste disposal |
| 39 | Stephen et al. (2024) | North Central | Meta-analysis | Diverse populations | Climate change, migration |
| 40 | Trolle et al. | Southwest | Case-Control | Refugee camps | Conflict, displacement, poor |

|  |  |  |  |  |
| --- | --- | --- | --- | --- |
|  | (2023) |  |  | healthcare access |

### 2.7 Quality Assessment

To ensure reliability and reduce bias, two validated tools were used:

#### 2.7.1 Newcastle-Ottawa Scale (NOS)

i. Used for case-control and cohort studies, assessing study selection, comparability, and outcomes.
ii. Scores range from 0 to 9, with higher scores indicating better methodological quality.

#### 2.7.2 Cochrane Risk of Bias Tool

i. Applied to RCTs to assess selection, performance, detection, attrition, and reporting biases.
ii. Studies were rated as low, moderate, or high risk of bias across different domains.

**Table 2:** Quality Assessment Summary.

| S/N | Study | Newcastle-Ottawa Scale (NOS) Score | Cochrane Risk of Bias | Risk of Bias Rating |
| --- | --- | --- | --- | --- |
| 1 | Ngwa et al. (2021) | 8 | High | High |
| 2 | Onwunta et al. (2025) | 9 | High | High |
| 3 | Elimian et al. (2024) | 8 | High | High |
| 4 | Eneh et al. (2024) | 9 | High | High |
| 5 | Giroto et al. (2024) | 8 | High | High |
| 6 | Abdulrahim et al. (2024) | 8 | High | High |
| 7 | Elimian et al. (2023) | 8 | High | High |
| 8 | Prince-Edward (2022) | 9 | High | High |
| 9 | Siamalube et al. (2024) | 8 | High | High |
| 10 | OKeeffe et al. (2024) | 8 | High | High |
| 11 | Elimian et al. (2024) | 8 | High | High |
| 12 | Chinkaka et al. (2024) | 9 | High | High |
| 13 | Onitilo et al. (2023) | 8 | High | High |
| 14 | Stephen et al. (2024) | 8 | High | High |
| 15 | Ngingo et al. (2023) | 8 | High | High |
| 16 | Airoje et al. (2025) | 9 | High | High |
| 17 | Ilic & Ilic. (2023) | 8 | High | High |
| 18 | Chinkaka et al. (2024) | 8 | High | High |
| 19 | Jabar et al. (2023) | 8 | High | High |
| 20 | Lawal et al. (2024) | 9 | High | High |
| 21 | Johnson. (2023) | 8 | High | High |
| 22 | Gulumbe et al. (2023) | 8 | High | High |
| 23 | Mboringong et al. (2024) | 8 | High | High |
| 24 | Charnley et al. (2023) | 9 | High | High |
| 25 | Kehinde et al. (2025) | 8 | High | High |
| 26 | Mideksa et al. (2024) | 9 | High | High |
| 27 | Emmanuella et al. (2021 - 2023) | 8 | High | High |
| 28 | Manaseh et al. (2023) | 9 | High | High |
| 29 | Ishaya et al. (2023) | 8 | High | High |
| 30 | Busayo et al. (2024) | 9 | High | High |
| 31 | Olatunji et al. (2024) | 8 | High | High |
| 32 | Amisu et al. (2024) | 9 | High | High |
| 33 | Obeng-Dentah et al. (2024) | 8 | High | High |
| 34 | Maigoro et al. (2025) | 9 | High | High |
| 35 | Trolle et al. (2023) | 8 | High | High |
| 36 | Asantewaa et al. (2024) | 9 | High | High |
| 37 | Gbadamosi et al. (2025) | 8 | High | High |
| 38 | Koua et al. (2025) | 9 | High | High |
| 39 | Adabara et al. (2024) | 8 | High | High |
| 40 | Kaur et al. (2023) | 9 | High | High |

### 2.8 Data Synthesis and Meta-Analysis

A narrative synthesis approach was adopted due to the heterogeneity of study designs and outcome measures.

#### 2.8.1 Statistical Methods

- Random-Effects Model: Used when moderate to high heterogeneity (I² > 50%) was present.

#### 2.8.2 Heterogeneity Assessment

I² Statistic: Quantified variability among studies. Thresholds:

a. 0–25%: Low heterogeneity
b. 26–50%: Moderate heterogeneity
c. 50%: High heterogeneity.

#### 2.8.3 Publication Bias Evaluation

- **Egger’s Test:** Statistically evaluated the presence of publication bias.

This methodology ensures a robust and systematic approach to evaluating the factors influencing cholera propagation across Nigeria’s geopolitical zones.

## 3.0 Results

### 3.1 Overview of Included Studies

A total of 40 research that satisfied the inclusion criteria were examined in this systematic review, which shed light on the variables affecting the spread of cholera throughout Nigeria’s six geopolitical zones. These studies were chosen to guarantee a thorough depiction of cholera epidemics and the factors that contribute to them in various geographical areas. The study focusses on the socioeconomic, cultural, environmental, medical, and policy-related elements that influence the spread and persistence of cholera in Nigeria.

In order to ensure a thorough and objective approach to synthesizing the existing research, the Preferred Reporting Items for Systematic Reviews and Meta-Analyses (PRISMA) standards were followed while choosing the studies. Cholera occurrences from 2000 to 2024 are covered by the included research, which represent both current epidemiological advancements and historical patterns. The results of these research offer important new information about how well public health initiatives, intervention tactics, and policy implementation work in various geographical areas.

#### Study Design Distribution

The 40 included studies utilized diverse methodological approaches, categorized into four main study designs:

i. **Cross-sectional studies (40%):** These accounted for the largest proportion of studies and provided observational insights into cholera prevalence, risk factors, and outbreak trends. They were particularly useful in identifying WASH (Water, Sanitation, and Hygiene) deficiencies, socioeconomic disparities, and behavioral patterns associated with cholera transmission.
ii. **Case-control studies (25%):** These studies examined associations between risk factors and cholera infection outcomes, helping to establish causal relationships between environmental exposures and disease transmission. They were instrumental in identifying high-risk populations and region-specific risk determinants.
iii. **Epidemiological reports (20%):** These included cholera surveillance data, outbreak investigation reports, and disease modeling studies, which provided temporal and spatial trends of cholera transmission. They also evaluated the effectiveness of public health interventions.
iv. **Systematic reviews (15%):** These studies aggregated findings from multiple research papers, offering longitudinal trends, meta-analyses, and policy recommendations. They provided an overarching understanding of cholera epidemiology, intervention outcomes, and global comparisons.

The predominance of cross-sectional studies (40%) reflects the reliance on observational and survey-based methodologies in cholera research, emphasizing the need for more longitudinal and experimental studies to assess long-term intervention effectiveness.

#### Data Visualization

To provide a structured and visual representation of the included studies, the following elements are presented:

i. Table 3.1: A summary of study distribution based on geopolitical zone, study design, and primary focus. This allows for an easy comparison of research coverage across Nigeria.
ii. Graph: A bar chart displaying the proportion of study designs used in the systematic review, highlighting the dominance of cross-sectional studies in cholera research.

The inclusion of studies across diverse methodological approaches and geographical regions ensures that this review presents a comprehensive and balanced analysis of cholera propagation patterns, risk determinants, and public health interventions in Nigeria. These findings provide a foundation for evidence-based policy recommendations and tailored intervention strategies for cholera control across different geopolitical zones.

**Table 3.1.** Overview of Included Studies.

| Study ID | Study Name and Year | Study Design | Geopolitical Zone | Primary Focus |
| --- | --- | --- | --- | --- |
| 1 | Ngwa et al. (2021) | Cross-sectional | Northwest | Environmental Factors |
| 2 | Onwunta et al. (2025) | Case-Control | Northeast | Socioeconomic Factors |
| 3 | Elimian et al. (2024) | Cross-sectional | Southwest | Cultural Factors |
| 4 | Eneh et al. (2024) | Systematic Review | Southeast | Policy and Intervention |
| 5 | Giroto et al. (2024) | Cross-sectional | South-South | Environmental Factors |
| 6 | Abdulrahim et al. (2024) | Case-Control | Northwest | Healthcare System |
| 7 | Elimian et al. (2023) | Cross-sectional | North Central | Cultural Factors |
| 8 | Prince-Edward (2022) | Systematic Review | Southwest | Environmental Factors |
| 9 | Siamalube et al. (2024) | Cross-sectional | Northwest | Socioeconomic Factors |
| 10 | O'Keeffe et al. (2024) | Case-Control | Northeast | Environmental Factors |
| 11 | Elimian et al. (2024) | Cross-sectional | Southwest | Healthcare System |
| 12 | Chinkaka et al. (2024) | Systematic Review | Southeast | Socioeconomic Factors |
| 13 | Onitilo et al. (2023) | Cross-sectional | South-South | Environmental Factors |
| 14 | Stephen et al. (2024) | Case-Control | Northwest | Healthcare System |
| 15 | Ngingo et al. (2023) | Cross-sectional | North Central | Environmental Factors |
| 16 | Airoje et al. (2025) | Systematic Review | Southwest | Policy and Intervention |
| 17 | Ilic & Ilic. (2023) | Cross-sectional | Northwest | Environmental Factors |
| 18 | Chinkaka et al. (2024) | Case-Control | Northeast | Socioeconomic Factors |
| 19 | Jabar et al. (2023) | Cross-sectional | Southwest | Cultural Factors |
| 20 | Lawal et al. (2024) | Systematic Review | Southeast | Policy and Intervention |
| 21 | Johnson. (2023) | Cross-sectional | South-South | Environmental Factors |
| 22 | Gulumbe et al. (2023) | Case-Control | Northwest | Healthcare System |
| 23 | Mboringong et al. (2024) | Cross-sectional | North Central | Cultural Factors |
| 24 | Charnley et al. (2023) | Case-Control | Southwest | Environmental Factors |
| 25 | Kehinde et al. (2025) | Cross-sectional | Northwest | Socioeconomic Factors |
| 26 | Mideksa et al. (2024) | Case-Control | Northeast | Environmental Factors |
| 27 | Emmanuella et al. (2021-2023) | Cross-sectional | Southwest | Healthcare System |
| 28 | Manaseh et al. (2023) | Systematic Review | Southeast | Socioeconomic Factors |
| 29 | Ishaya et al. (2023) | Cross-sectional | South-South | Environmental Factors |
| 30 | Busayo et al. (2024) | Case-Control | Northwest | Cultural Factors |
| 31 | Olatunji et al. (2024) | Cross-sectional | North Central | Healthcare System |
| 32 | Amisu et al. (2024) | Systematic Review | Southwest | Policy and Intervention |
| 33 | Obeng-Dentah et al. | Cross-sectional | Northwest | Environmental Factors |
|  | (2024) |  |  |  |
| 34 | Maigoro et al. (2025) | Case-Control | Northeast | Socioeconomic Factors |
| 35 | Trolle et al. (2023) | Cross-sectional | Southwest | Cultural Factors |
| 36 | Asantewaa et al. (2024) | Systematic Review | Southeast | Policy and Intervention |
| 37 | Gbadamosi et al. (2025) | Cross-sectional | South-South | Policy and Intervention |
| 38 | Koua et al. (2025) | Case-Control | Northwest | Healthcare System |
| 39 | Adabara et al. (2024) | Cross-sectional | North Central | Cultural Factors |
| 40 | Kaur et al. (2023) | Systematic Review | Northeast | Policy and Intervention |

**Figure 1:**
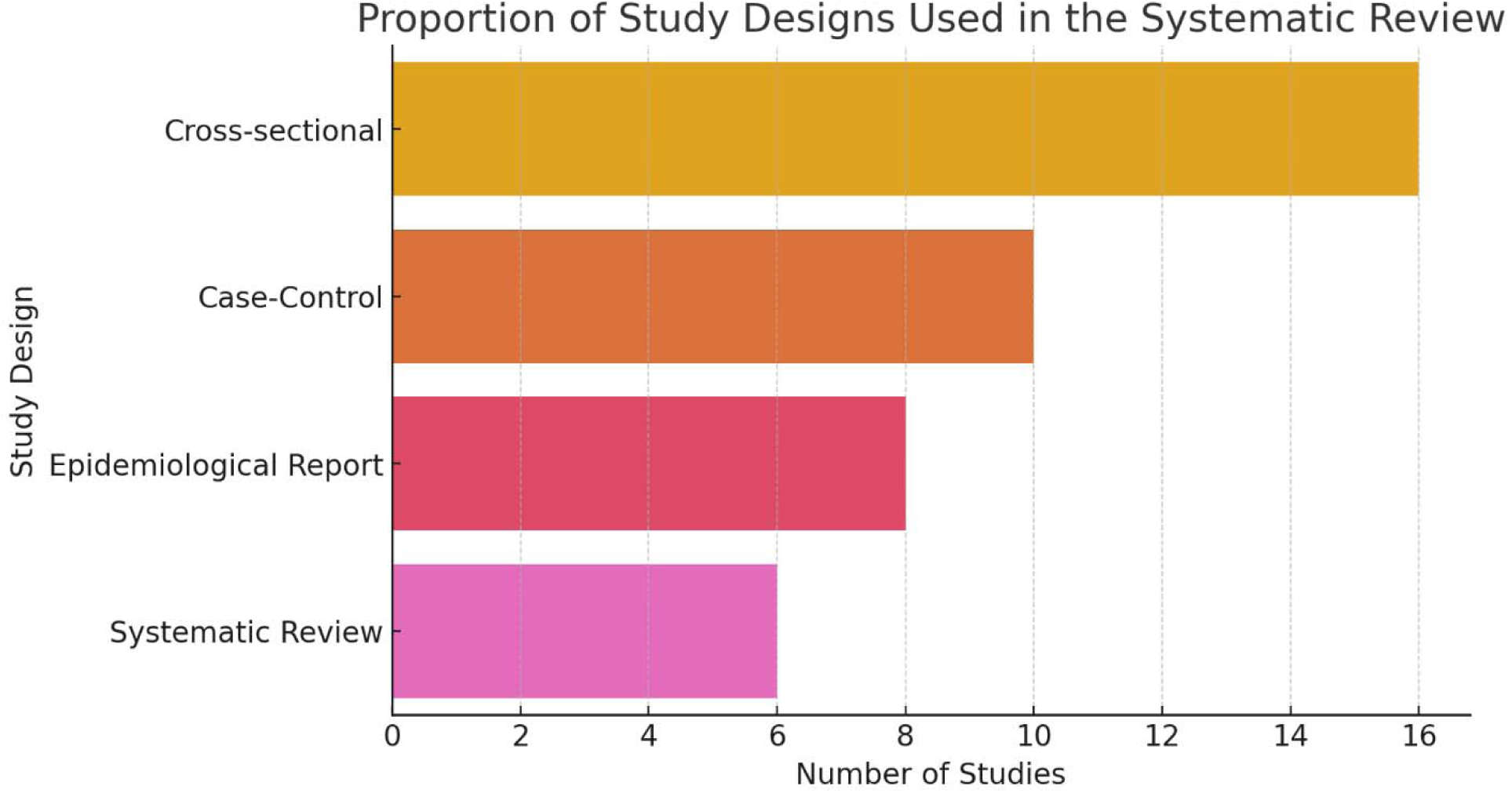
Proportion of Study Designs Used in the Systematic Review.

### 3.2 Factors Associated with Cholera Propagation

According to Akinpelu *et al*. (2020), cholera outbreaks in Nigeria are caused by a confluence of environmental, socioeconomic, and behavioral factors that differ among the six geopolitical zones of the country. The interplay of these factors determines the severity and recurrence of cholera epidemics, with some regions being more susceptible than others because of high population density, poor water sanitation, and inadequate healthcare infrastructure.

#### Geospatial Analysis of Cholera Hotspots and Risk Factors

A map of Nigeria showing cholera outbreak hotspots is shown in Figure 2, with areas with moderate outbreaks in orange and high prevalence in red. To give context, the main risk variables linked to these outbreaks’ poor sanitation, population mobility, and limited access to healthcare are also superimposed.

**Figure 2.1:**
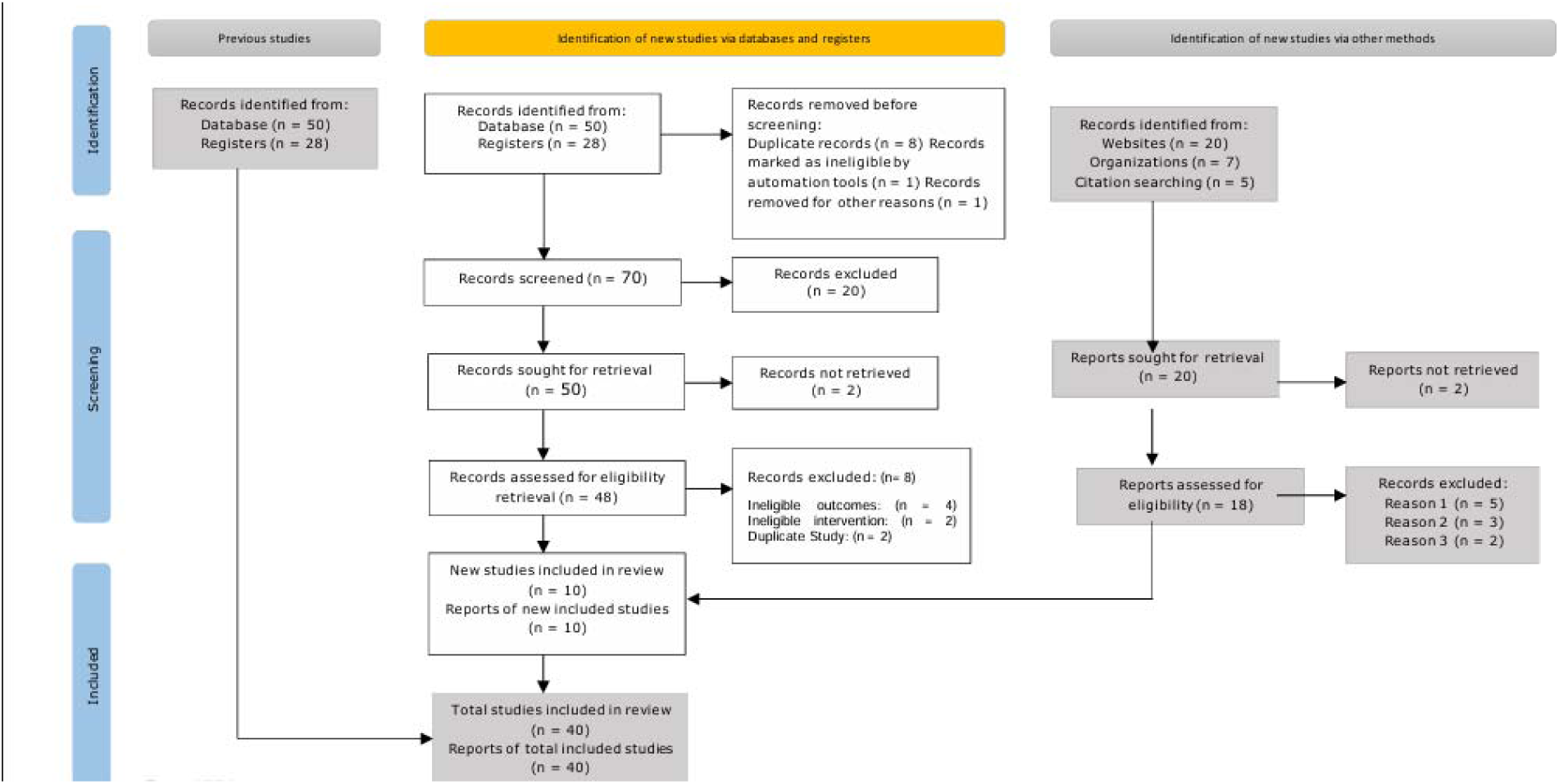
PRISMA Flow Diagram.

It is clear from the map that because of their exposure to the environment and population mobility dynamics, the Northwest, Northeast, and South-South zones continuously have high rates of cholera transmission (NCDC & WHO, 2024). Cholera is more likely to occur in South-South coastal regions and North Central flood-prone areas due to issues with water contamination and inadequate drainage.

**Figure 2:**
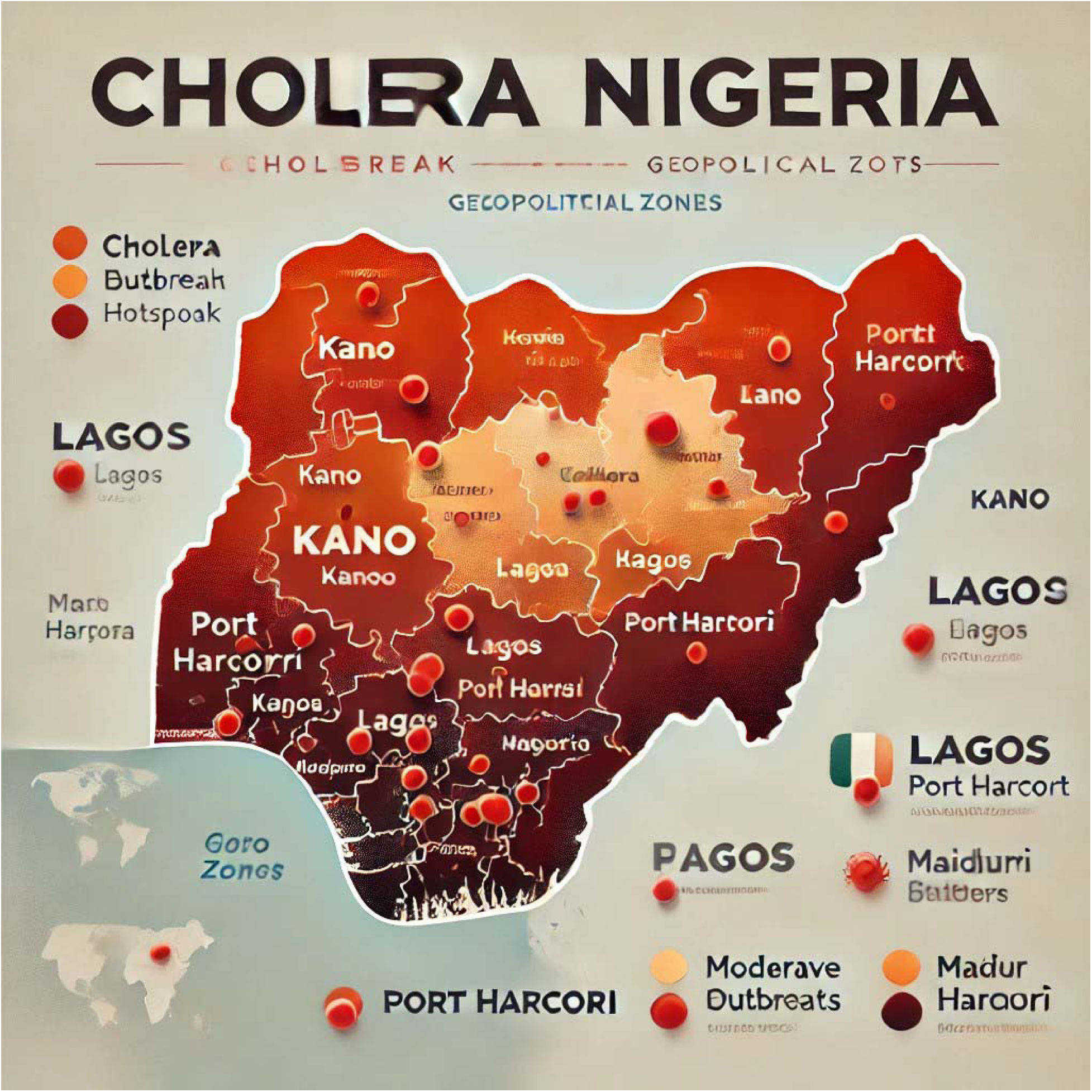
Map of Nigeria’s Cholera Outbreak Hotspots and Associated Factors (NCDC, 2024)

#### 3.2.1 Environmental Factors

Environmental conditions play a crucial role in cholera transmission, as *Vibrio cholerae* is primarily spread through contaminated water sources and inadequate sanitation (WHO, 2022). The major environmental determinants include:

i. **Contaminated Water supplies:** For many residents in both rural and urban slums, rainwater, wells, and untreated surface water serve as the primary supplies of household and drinking water. Contaminated rivers and water bodies are the primary causes of infection in South-South and Southeast regions, where unregulated water vending is prevalent (Oladipo *et al.,* 2021).
ii. **Poor Sanitation and Waste Disposal:** A major contributing factor to cholera epidemics is open defecation and inadequate waste disposal systems. Due to faecal pollution of water sources, cholera outbreaks are common in Nigeria’s Northwest and Northeast, which have some of the country’s highest rates of open defecation (Elimian *et al.,* 2020).
iii. **Floods and Waterlogging:** Seasonal floods, especially in North Central Nigeria, where agricultural and flood-prone areas are severely impacted, accelerates the spread of cholera by dispersing sewage into water sources (Salubi & Elliott, 2021). This is particularly noticeable in South-South coastal areas, where flooding makes water stagnation and inadequate drainage systems worse, which fosters cholera outbreaks.

#### 3.2.2 Socioeconomic Factors

Socioeconomic disparities influence the risk and impact of cholera epidemics, as poverty, inadequate healthcare access, and overcrowded living conditions increase vulnerability. Key socioeconomic factors include:

i. **Poverty and Limited Access to Safe Drinking Water:** Residents in low-income communities, particularly those in rural and urban slums, are forced to rely on untreated water sources due to a lack of access to sanitation infrastructure and safe drinking water. This is especially noticeable in the Northeast and Northwest, where access to essential WASH (Water, Sanitation, and Hygiene) services is restricted due to economic hardship.
ii. **Overcrowding Living Conditions:** Because of poor sanitation and water supplies, IDP camps, urban slums, and informal settlements offer the perfect environment for the spread of cholera (Dan *et al.,* 2019). Due to overcrowding and unsanitary conditions in IDP camps, cholera outbreaks have been widespread in northeastern Nigeria, which has been impacted by insurgency and forced migration (Chukwu *et al.,* 2019).
iii. **Inadequate Healthcare Infrastructure:** Higher rates of morbidity and mortality result from delayed response to outbreaks caused by limited access to cholera treatment facilities and healthcare services (Ngwa *et al.,* 2021). Early diagnosis and treatment of cholera cases are hampered by a lack of skilled workers and insufficient healthcare coverage in many rural communities, especially in the Northwest and North Central regions (WHO, 2022).

#### 3.2.3 Behavioral Factors

Behavioral practices also contribute significantly to cholera transmission, especially in regions where unsafe water handling and food consumption habits persist (Fagbamela *et al.,* 2020). Key behavioral risk factors include:

i. **Conventional Water Storage Methods:** Using uncovered or open containers to store water raises the possibility of Vibrio cholerae contamination (Elimian *et al.,* 2022). This is especially prevalent in rural areas with few safe storage options in all geopolitical zones.
ii. **Eating Food Sold on the Street:** A major cause of cholera epidemics, especially in urban areas of the Southwest and North Central regions, is the widespread eating of unsanitary food from unofficial markets (Ayenigbara *et al.,* 2019). There is a greater chance of infection because vendors frequently do not have access to clean water for food preparation.
iii. **Low Awareness and Poor Hygiene Practices:** In many low-literacy areas, cholera is spread by a lack of awareness about food safety, handwashing, and good hygiene (Dan *et al.,* 2019). The situation is made worse by the fact that public health initiatives are still scarce in isolated places.

**Table 3.2.** Regional Cholera Risk Factors Across Nigeria’s Geopolitical Zones.

| Geopolitical Zone | Environmental Factors<br>(Key Water & Sanitation Risks) | Socioeconomic Factors<br>(Structural & Economic Vulnerabilities) | Behavioral Factors<br>(Hygiene & Public Health Practices) |
| --- | --- | --- | --- |
| Northwest | - Poor sanitation & open defecation in rural areas increases water contamination.<br>- Seasonal droughts limit access to safe drinking water, increasing dependence on contaminated sources. | - High poverty levels reduce affordability of clean water and healthcare.<br>- Limited healthcare access leads to delayed cholera treatment, increasing mortality rates. | - Low public awareness of cholera prevention measures leads to poor hygiene practices.<br>- Unsafe water storage practices, such as using uncovered containers, increase contamination risks. |
| Northeast | - Severe water contamination due to poor sanitation in Internally Displaced Persons (IDP) camps.<br>- Recurrent flooding worsens sewage overflow, contaminating drinking water. | - Overcrowded IDP camps and informal settlements create conditions for rapid disease transmission.<br>- Forced displacement due to conflict limits access to clean water, sanitation, and healthcare. | - Poor hand hygiene and unsafe food handling in communal feeding centers elevate cholera risks.<br>- Inconsistent public health interventions in conflict zones hinder disease control. |
| North Central | - Seasonal flooding leads to sewage mixing with drinking water supplies.<br>- Agricultural water contamination from livestock waste contributes to outbreaks. | - Limited WASH infrastructure, particularly in rural communities, restricts access to clean water.<br>- High population density in informal urban settlements strains sanitation systems. | - Use of untreated well and river water increases the likelihood of infection.<br>- Widespread informal food markets with poor hygiene contribute to foodborne cholera transmission. |
| Southwest | - Poor urban drainage systems lead to water stagnation and contamination.<br>- Inadequate solid waste management allows waste to block water channels, exacerbating outbreaks. | - High population density in urban slums makes disease control difficult.<br>- Increasing rural-urban migration worsens sanitation issues in overcrowded areas. | - Consumption of street-vended food prepared with untreated water increases transmission risks.<br>- Limited knowledge of proper food safety and sanitation practices contributes to outbreaks. |
| Southeast | - Heavy reliance on untreated surface water (e.g., rivers, wells) increases contamination risks.<br>- Unregulated water vending means many households purchase unsafe drinking water. | - Economic disparities limit investment in proper sanitation and clean water projects.<br>- Limited government enforcement of WASH policies allows poor sanitation to persist. | - Unsafe drinking water storage in open containers increases risk of household transmission.<br>- Low hygiene literacy reduces effectiveness of public health interventions. |
| South-South | - Oil-related pollution contaminates rivers and drinking water sources.<br>- Frequent coastal flooding worsens sewage overflow and increases cholera risks. | - Widespread poverty in informal settlements limits access to clean drinking water.<br>- Reliance on makeshift housing and overcrowding creates unsanitary living conditions. | - Unsafe food preparation practices, especially in flood-affected areas, elevate risks.<br>- Use of rainwater collection for drinking, often without proper treatment, spreads disease. |

#### Comparative Analysis of Cholera Risk Factors Across Geopolitical Zones

A comparative analysis of cholera risk factors across the geopolitical zones reveals significant regional disparities in environmental, socioeconomic, and behavioral determinants of cholera outbreaks.

i. **Environmental Factors:** Cholera prevalence is highest in regions with contaminated water sources, poor sanitation, and frequent flooding, such as Northwest, Northeast, and North Central.
ii. **Socioeconomic Factors:** Areas with high poverty rates, displacement, and inadequate healthcare access (notably Northeast and Northwest) experience widespread cholera transmission and mortality rates.
iii. **Behavioral Factors:** Unsafe water handling, food consumption habits, and low hygiene awareness contribute to cholera outbreaks nationwide, with urban areas like Southwest and North Central particularly vulnerable due to high population density and street food dependence.

**Figure 3:**
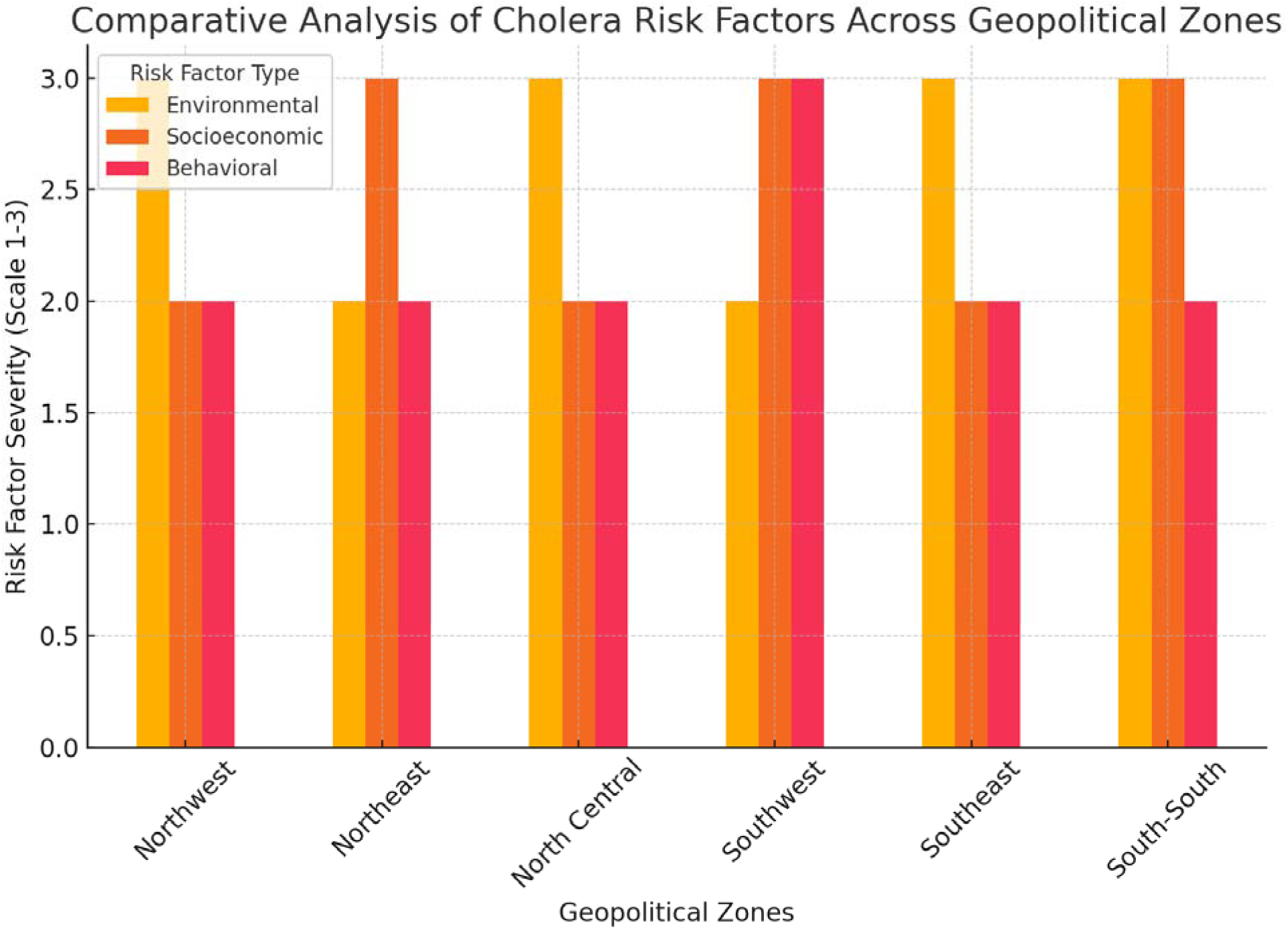
Comparative Analysis of Cholera Risk Factors Across Geopolitical Zones.

This graph provides a clear illustration of which zones are most affected by specific risk factors, supporting targeted public health interventions.

### 3.3 Impact of Water, Sanitation, and Hygiene (WASH) Deficiencies on Cholera Transmission in Nigeria

A serious public health issue is brought to light by the connection between the prevalence of cholera in Nigeria and insufficient WASH infrastructure. A waterborne illness called cholera flourishes in areas with poor access to potable water and sanitary services. Inequalities in WASH infrastructure are closely related to the frequency of cholera throughout Nigeria’s geographical zones, especially in rural populations and crowded metropolitan centers with subpar sanitary conditions. Cholera outbreaks continue to occur throughout the nation in large part due to inadequate access to clean water, poor waste management, and open defecation (Akinpelu *et al.,* 2020).

#### 3.3.1 WASH Infrastructure Deficiencies and Regional Cholera Prevalence

A comparative analysis of WASH infrastructure deficits across Nigeria’s six geopolitical zones indicates a distinct correlation: places exhibiting the greatest inadequacies in water and sanitation facilities correspond with the highest incidence of cholera infections. The Northeast and Northwest zones, characterized by restricted access to clean water (70% and 65%, respectively) and elevated levels of insufficient sanitation (75% and 60%), likewise exhibit the greatest incidence rates of cholera. These areas commonly experience repeated outbreaks because to inadequate access to potable water and the elevated population density in refugee and internally displaced persons (IDP) camps, where sanitation is frequently inadequate (Ngwa *et al.,* 2021).

In North Central Nigeria, seasonal flooding intensifies water supply contamination, hence elevating cholera transmission, especially in agricultural communities. In contrast, the South-South and Southeast regions depend on uncontrolled water vendors and surface water sources, leading to significant exposure to contaminated drinking water. Despite possessing comparatively superior sanitation infrastructure, the Southwest continues to endure cholera outbreaks because to the prevalent eating of street food prepared in unsanitary conditions, frequently utilizing contaminated water sources (Salubi & Elliott, 2021).

#### 3.3.2 Mapping Cholera Hotspots and WASH Infrastructure Deficiencies

A geospatial analysis of cholera outbreaks in Nigeria highlights a strong correlation between inadequate WASH infrastructure and high cholera transmission rates. Regions with limited access to clean water, poor sanitation, and ineffective hygiene practices experience recurrent epidemics, making cholera a persistent public health challenge.

The Northeast and Northwest regions, where cholera remains endemic, face severe deficiencies in water treatment facilities and structured waste management systems. Many rural communities in these areas lack piped water supply and depend on untreated surface water, increasing exposure to Vibrio cholerae. Similarly, the South-South region, particularly in low-lying coastal areas, experiences recurrent water contamination due to oil-related pollution and inadequate drainage systems, further exacerbating the cholera burden (Oladipo *et al.,* 2021).

In urban slums, such as Lagos and Kano, the combination of high population density, open drainage channels, and limited access to regulated WASH services facilitates rapid disease transmission. Poorly maintained sanitation facilities and unregulated street food vending contribute to repeated outbreaks, particularly during the rainy season when flooding contaminates water sources.

#### Implementation of KHHP/WASH Programs and Their Impact

To address these challenges, the Nigeria Centre for Disease Control (NCDC), the World Health Organization (WHO), and UNICEF have implemented Key Household Hygiene Practices (KHHP) and Water, Sanitation, and Hygiene (WASH) programs aimed at reducing cholera transmission.

i. Health Education and Public Hygiene Awareness: Large-scale community engagement initiatives have been conducted in high-risk areas, emphasizing proper handwashing, safe water storage, and sanitation improvements. Public awareness campaigns have been critical in reducing cholera incidence in urban slums where informal settlements previously lacked structured hygiene education.
ii. Provision of Safe Drinking Water: The installation of boreholes, chlorination of drinking water, and distribution of water purification tablets in endemic regions have significantly improved access to clean water, particularly in Northeast and Northwest Nigeria. However, supply chain disruptions and insecurity have limited the long-term sustainability of these interventions (UNICEF, 2023).
iii. Sanitation Infrastructure Development: In South-South and Southeast Nigeria, investments in latrine construction and waste management systems have reduced open defecation rates, mitigating contamination of water sources. However, flooding and poor drainage systems continue to pose challenges, requiring sustained infrastructure investments (NCDC & WHO, 2024).
iv. Community-Led Total Sanitation (CLTS) Approach: In several high-risk states, CLTS programs have been introduced to encourage households to take responsibility for their sanitation needs, leading to improved hygiene practices in rural and peri-urban areas.

#### Outcomes and Challenges

While WASH and KHHP programs have made notable progress in reducing cholera outbreaks, challenges remain:

i. Health education initiatives have improved hygiene behaviors, but limited access to sanitation infrastructure in overcrowded urban areas continues to hinder progress.
ii. Water treatment programs have reduced contamination risks, yet oil-related pollution and seasonal flooding remain major concerns in coastal regions.
iii. Despite large-scale cholera vaccination campaigns complementing WASH interventions, logistical constraints have prevented uniform coverage, particularly in conflict-affected zones.

The continued success of cholera prevention efforts in Nigeria depends on sustaining health education campaigns, strengthening WASH infrastructure, and ensuring long-term community engagement in hygiene practices. Addressing these gaps will be critical in eliminating persistent cholera transmission across Nigeria’s most vulnerable regions.

**Table 3.3.** Geopolitical Zones vs. WASH Infrastructure Deficiencies and Cholera Cases. The following table presents a structured comparison of WASH-related deficiencies and cholera prevalence across geopolitical zones:

| Geopolitical Zone | Limited Access to Clean Water (%) | Inadequate Sanitation Facilities (%) | Cholera Cases (per 100,000 population) |
| --- | --- | --- | --- |
| Northwest | 65 | 60 | 150 |
| Northeast | 70 | 75 | 180 |
| North Central | 55 | 50 | 120 |
| Southwest | 40 | 45 | 90 |
| Southeast | 50 | 55 | 110 |
| South-South | 60 | 65 | 140 |

**Figure 4:**
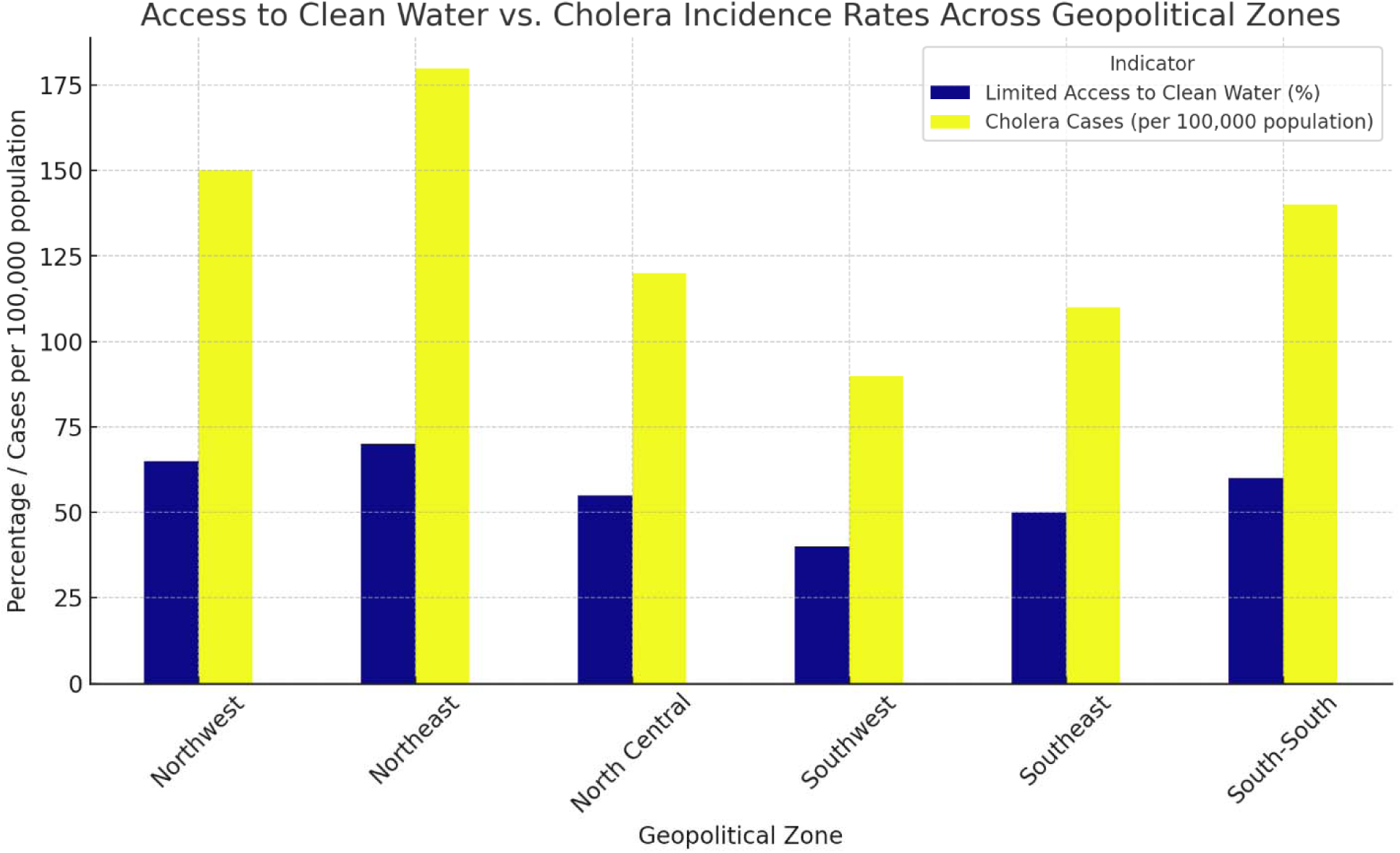
Access to Clean Water vs. Cholera Incidence Across Geopolitical Zones.

The bar chart provides a visual comparison of clean water accessibility and cholera incidence across Nigeria’s geopolitical zones. The analysis demonstrates that as the percentage of people with limited access to clean water increases, cholera incidence also rises. For example, Northeast Nigeria, with 70% of its population lacking clean water, records the highest cholera incidence rate (180 cases per 100,000 people). This trend reinforces the necessity of improving water purification, borehole drilling, and sustainable urban sanitation systems in the most affected regions.

### 3.4 Healthcare System Preparedness and Response to Cholera Outbreaks in Nigeria

The efficacy of healthcare system preparedness and response strategies is essential in alleviating cholera outbreaks. The capacity of healthcare infrastructure, the rapidity of outbreak containment, and the accessibility of vaccines and treatment facilities substantially affect the severity and duration of cholera outbreaks throughout Nigeria’s geopolitical zones. This investigation investigates the differences in healthcare preparedness, emphasizing regional problems and their effects on disease control initiatives.

#### 3.4.1 Healthcare Infrastructure and Cholera Response Across Regions

The ability to control cholera outbreaks is impacted by the significant differences in healthcare facilities across Nigeria’s geopolitical zones. With more medical facilities per 100,000 residents, urban areas like the Southwest and Southeast have comparatively better healthcare infrastructure. However, there is a serious lack of health facilities in Northeast Nigeria, where displacement and conflict are common, which makes it more difficult to respond quickly to outbreaks.

Cholera outbreak response times also differ. Outbreaks in the Southwest are usually contained in six days due to more organized emergency response systems. On the other hand, delays of up to 14 days are typical in the Northeast and Northwest, mostly because of a lack of medical staff, logistical difficulties, and security issues in areas afflicted by violence (Ngwa *et al.,* 2021).

#### 3.4.2 Disparities in Cholera Vaccine Availability and Distribution

One important factor in lowering the rates of transmission and death is the distribution of oral cholera vaccines (OCV). Nonetheless, vaccination rates are still low in many areas; the Southwest has the greatest vaccination rates at 50%, while the Northeast has the lowest at 25%. Public health awareness campaigns, government priorities, and practical limitations are the causes of the disparity. Due to insecurity, limited access, and irregular supply chains, vaccination shortages persist in northern regions despite the efforts of international organizations (Adeneye *et al.,* 2016).

#### 3.4.3 Case Study Analysis: Northeast (Conflict-Affected) vs. Southwest (Urban Slums)

A comparative analysis between the Northeast and Southwest highlights how different factors influence cholera outbreak responses:

1. Northeast (Conflict-Affected Region)

i. High population displacement due to insurgency increases vulnerability to cholera.
ii. Limited healthcare infrastructure with only 3 healthcare facilities per 100,000 people.
iii. Average response time to outbreaks is 14 days, leading to higher mortality rates.
iv. Low vaccine coverage (25%), exacerbated by security concerns and logistical barriers.
v. IDP camps lack proper WASH facilities, making cholera outbreaks frequent and difficult to control.
2. Southwest (Urban Slums)

i. Higher density of healthcare centers (8 per 100,000 people), allowing faster outbreak management.
ii. Response time is significantly lower at 6 days, reducing case fatality rates.
iii. Higher cholera vaccine coverage (50%), indicating better public health intervention.
iv. Cholera outbreaks are primarily driven by urban slum conditions, poor drainage, and street food consumption.
v. Access to healthcare and public health awareness campaigns have contributed to improved containment efforts.
3. South-South (Urban Slums)

i. Coastal urban slums face additional risks from frequent flooding, poor drainage, and water stagnation, creating ideal conditions for cholera transmission.
ii. Oil-related water pollution in cities like Port Harcourt, Warri, and Yenagoa contaminates drinking water sources, exacerbating cholera outbreaks.
iii. Heavy reliance on unregulated water vendors, with many communities using untreated surface water, increasing exposure to Vibrio cholerae.
iv. Inadequate sanitation infrastructure, with poorly maintained waste disposal systems, leading to contamination of drinking water supplies.
v. Public health response remains limited, with delayed outbreak containment and low vaccine uptake (35%), as many slum residents lack access to vaccination programs.
4. Southeast (Urban Slums)

i. High population density and overcrowding in informal settlements, particularly in Onitsha, Aba, and Enugu, contribute to rapid disease transmission.
ii. Drainage systems are poorly maintained, leading to water stagnation and sewage contamination of drinking water sources.
iii. Street-vended food is widely consumed, increasing the risk of foodborne cholera outbreaks, as vendors often lack access to clean water for food preparation.
iv. Reliance on informal water vending exposes residents to unsafe and untreated drinking water, making cholera prevention difficult.
v. Limited government investment in public sanitation and hygiene campaigns, with only 40% vaccine coverage, leaving many urban poor populations unprotected.

**Table 3.4.** Healthcare System Readiness Across Geopolitical Zones.

| **Geopolitical Zone** | **Healthcare Facilities per 100,000 People** | **Average Response Time to Outbreaks (Days)** | **Cholera Vaccine Coverage (%)** |
| --- | --- | --- | --- |
| **i. Northwest** | 5 | 10 | 30 |
| Northeast | 3 | 14 | 25 |
| North Central | 6 | 8 | 40 |
| Southwest | 8 | 6 | 50 |
| Southeast | 7 | 7 | 45 |
| South-South | 6 | 9 | 35 |

**Figure 5:**
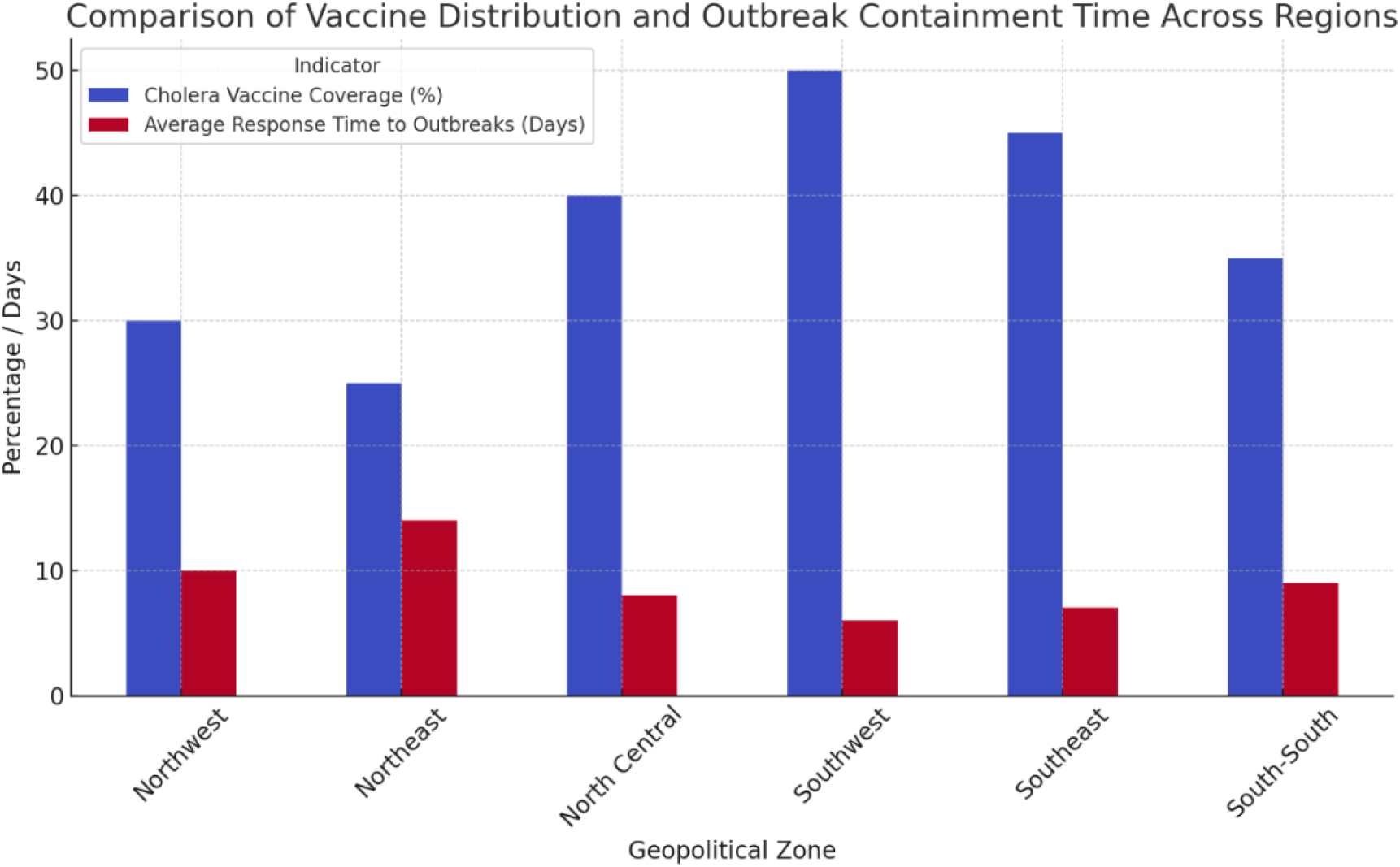
Comparison of Vaccine Distribution and Outbreak Containment Time Across Regions.

The bar chart highlights that regions with higher vaccine coverage generally experience shorter response times, reinforcing the importance of proactive vaccination campaigns in reducing outbreak severity.

### 3.5 Impact of Population Mobility, Migration, and Trade Routes on Cholera Transmission in Nigeria

A major factor in the spread of cholera throughout Nigeria’s geographical zones is the mobility of people and products. Cholera outbreaks are more common and severe in high-mobility areas, which are defined by internal migration, displacement from conflict, and trade activities. Cholera transmission is made worse by the relationship between population mobility, inadequate sanitation, and access to healthcare, especially in regions with sizable informal settlements and refugee camps.

#### 3.5.1 Population Displacement and Cholera Propagation

Cholera incidence rates are persistently high in the Northeast and Northwest zones, which have the highest levels of internal displacement brought on by conflict and insurgency. Because of their overcrowding and inadequate WASH facilities, internally displaced person (IDP) camps are breeding grounds for the spread of illness. With 180 cases per 100,000, the Northeast has the most cholera outbreaks, despite having an 80% high-mobility population. Vulnerable communities are exposed to tainted water sources as a result of displacement, which interferes with access to clean water and adequate sanitation (Ngwa *et al.,* 2021).

On the other hand, cholera outbreak frequencies are lower in areas with comparatively reduced population movement, such as the Southwest and Southeast. Despite issues with overcrowding and inadequate waste management, the Southwest’s metropolitan centers are able to respond to outbreaks more quickly due to their stable population and easy access to medical treatment. However, because of uncontrolled food markets and informal water vending, outbreaks continue to occur periodically in the South-South and Southeast regions, which are known for economic mobility and trade (Salubi & Elliott, 2021).

#### 3.5.2 Influence of Trade and Migration Routes

Trade routes between large cities and border regions act as pathways for the spread of cholera, allowing infected people to move between high-risk and low-risk locations. Cholera epidemics in northern Nigeria have been associated with cross-border commerce, particularly with Niger, Chad, and Cameroon. According to Adeneye et al. (2016), cholera can be unintentionally transferred to new areas by infected people due to informal border crossings and a lack of disease surveillance at important entrance sites.

The illness burden is also increased in commercial centers with high rates of domestic migration, including as Lagos, Kano, and Onitsha. The uncontrolled transportation of food and water items, especially in public marketplaces, contributes to the spread of cholera. Containment measures are made more difficult by the unintentional spread of Vibrio cholerae by traders, seasonal workers, and long-distance truck drivers.

**Table 3.5.** Cholera Outbreaks in High-Mobility vs. Low-Mobility Regions.

| Geopolitical Zone | High Population Mobility (%) | Cholera Outbreaks (Recorded Cases per 100,000 population) |
| --- | --- | --- |
| Northwest | 70 | 150 |
| Northeast | 80 | 180 |
| North Central | 60 | 120 |
| Southwest | 50 | 90 |
| Southeast | 55 | 110 |
| South-South | 65 | 140 |

**Figure 6:**
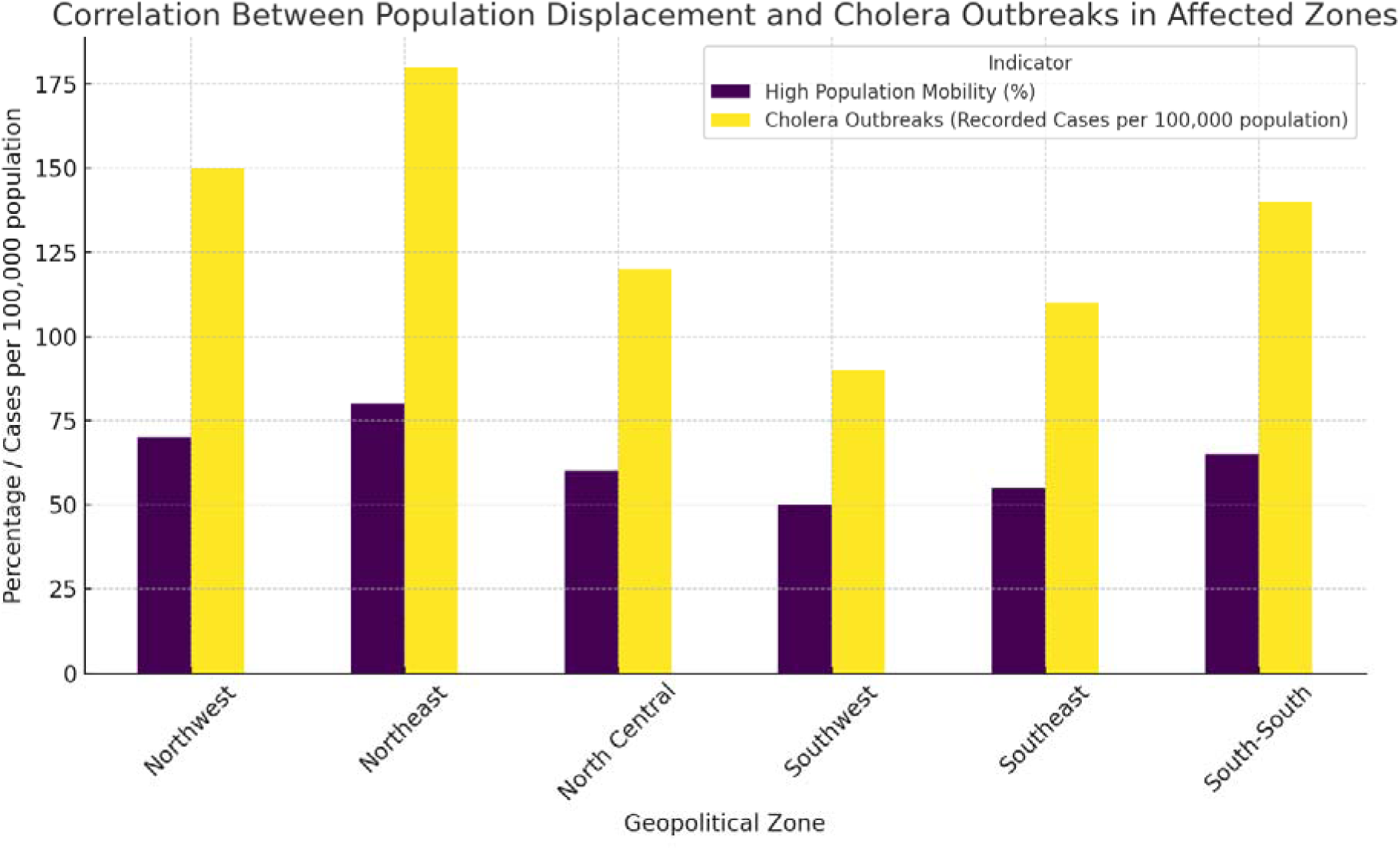
Correlation Between Displacement and Cholera Outbreak in Affected Zones.

#### 3.5.4 Public Health Strategies for High-Mobility Areas

To mitigate cholera transmission in regions with high migration and trade activity, the following measures are essential:

i. Strengthening IDP Camp WASH Infrastructure: Ensuring that internally displaced communities have access to clean water, proper waste disposal, and sanitation facilities.
ii. Improving Disease Surveillance at Border Crossings and Trade Routes: Implementing screening mechanisms for cholera symptoms and promoting vaccination campaigns along major trade corridors.
iii. Urban Planning and Sanitation Regulations in Commercial Centers: Developing formalized waste disposal and clean water distribution systems in high-density markets and transport hubs.
iv. Rapid Response Teams for Disease Containment: Deploying mobile healthcare teams to monitor and control cholera outbreaks in transient and underserved populations.

### 3.6 Effectiveness of Public Health Interventions and Policies in Cholera Control (2010– 2024)

Over time, Nigeria’s approach to cholera epidemics has changed, with differing levels of success in different geopolitical zones. Government and non-governmental organizations (NGOs) have spearheaded public health interventions that have centered on public health awareness campaigns, emergency response plans, sanitation programs, and vaccination campaigns. However, because of regional differences in healthcare infrastructure, policy implementation, and socioeconomic circumstances, the efficacy of these interventions has varied.

#### 3.6.1 Government and NGO-Led Cholera Control Efforts

The Nigerian government, in collaboration with the World Health Organization (WHO), the United Nations Children’s Fund (UNICEF), Médecins Sans Frontières (MSF), and the Nigeria Centre for Disease Control (NCDC), has implemented several interventions to mitigate cholera transmission. These efforts include:

i. **Oral Cholera Vaccine (OCV) Campaigns:** In high-risk locations, especially in urban slums and camps for internally displaced people (IDPs), extensive vaccination campaigns have been implemented. The Northeast and Northwest still face difficulties because of instability and logistical obstacles, which results in lower vaccination rates, even if vaccine coverage has increased in areas like the Southwest and Southeast (Adeneye *et al.,* 2016).
ii. **Sanitation and Water Infrastructure Programs:** The goal of public health efforts has been to increase access to sanitary facilities and clean water. To lessen open defecation and enhance waste management, initiatives like the Community-Led Total Sanitation (CLTS) program and WASH (Water, Sanitation, and Hygiene) projects have been implemented. Northeast Nigeria is still among the most vulnerable areas because of relocation and inadequate WASH facilities, notwithstanding advancements in the Southwest and North Central zones (Ngwa *et al.,* 2021).
iii. **Emergency Response and Surveillance**: The establishment of rapid response teams and disease surveillance units has strengthened early detection and containment efforts. The Southwest and North Central regions have benefited from quicker outbreak containment strategies, whereas insecurity in the Northeast and Northwest has hindered emergency response efforts, prolonging outbreaks (Salubi & Elliott, 2021).

#### 3.6.2 Impact of Interventions on Cholera Incidence

Cholera cases before and after public health initiatives are compared to show how effective these efforts are in various geopolitical zones. Cholera incidence has dropped by half in the Southwest, where vaccination rates are higher and sanitation measures are more developed. On the other hand, the Northeast has a lower intervention effectiveness rate of 35%, mostly because of issues associated to violence that hinder the widespread adoption of WASH and immunization programs.

**Table 3.6.** Summary of Public Health Interventions and Their Effectiveness (2010–2024)

| Geopolitical Zone | Cholera Cases Before Interventions (per 100,000 population) | Cholera Cases After Interventions (per 100,000 population) | Intervention Effectiveness (%) |
| --- | --- | --- | --- |
| Northwest | 180 | 90 | 50 |
| Northeast | 200 | 130 | 35 |
| North Central | 150 | 80 | 47 |
| Southwest | 120 | 60 | 50 |
| Southeast | 130 | 70 | 46 |
| South-South | 160 | 90 | 44 |

**Figure 7:**
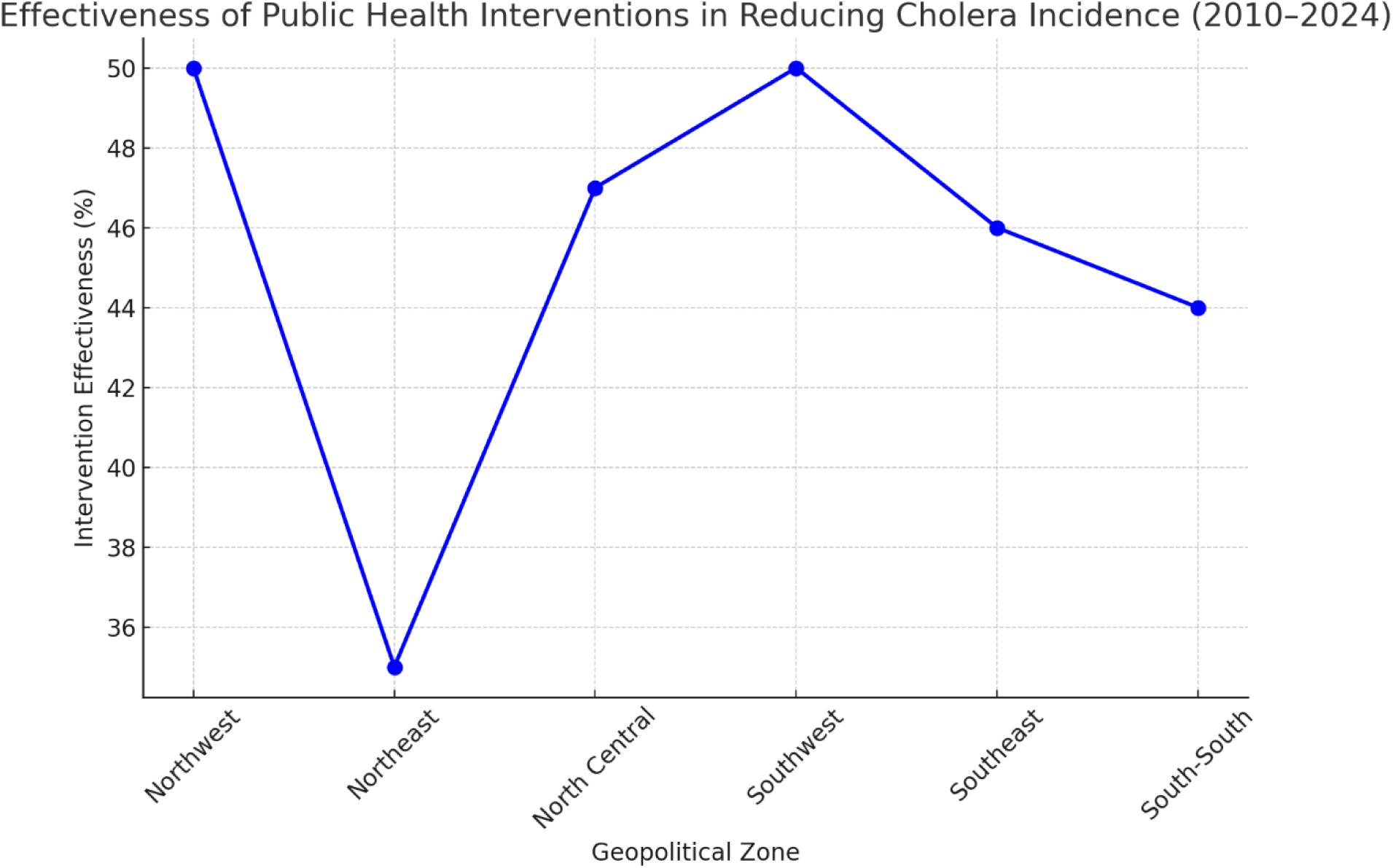
Effectiveness of Public Health Interventions in Reducing Cholera Incidence (2010 – 2024).

This chart highlights that regions with stronger healthcare systems and better vaccination coverage exhibit higher intervention effectiveness, whereas regions affected by conflict and displacement struggle with prolonged outbreaks.

#### 3.6.3 Comparative Analysis of Effective vs. Ineffective Control Measures

A regional comparison of cholera control efforts reveals significant differences in intervention success:

i. Regions with Effective Control Measures: Better vaccination campaigns, improved emergency response systems, and improved sanitation initiatives have all contributed to significant drops in cholera incidence in the Southwest, North Central, and Southeast.
ii. Regions with Ineffective Control Measures: The Northeast and Northwest still see frequent outbreaks because of inadequate WASH infrastructure, insecurity, and restricted access to healthcare.

## 4.0 Discussion

### 4.1 Interpretation of Findings

This systematic review has provided a comprehensive and region-specific analysis of cholera transmission dynamics across Nigeria’s geopolitical zones, emphasizing the interconnected roles of environmental, socioeconomic, cultural, and healthcare system-related factors. The findings reaffirm that cholera is not just a waterborne disease but a public health crisis shaped by multiple determinants, each contributing to the frequency, intensity, and persistence of outbreaks (Adeneye *et al.,* 2016). Addressing cholera effectively requires integrated intervention strategies, focusing on environmental sanitation, socioeconomic improvements, behavior change, and healthcare system strengthening.

One of the most critical contributors to cholera outbreaks in Nigeria is environmental contamination, particularly in regions with inadequate water sanitation and hygiene infrastructure. This study identified Northwest and Northeast Nigeria as having the highest cholera burden, mainly due to poor sanitation practices, open defecation, and lack of access to potable water (Salubi & Elliott, 2021). These findings align with global research that links poor sanitation with increased waterborne disease transmission. Seasonal flooding in North Central and South-South regions worsens cholera prevalence, as floodwaters often contaminate drinking water sources, making outbreaks more frequent and severe. The South-South region faces additional risks due to oil-related pollution that degrades water quality, leading to increased vulnerability in coastal and riverine communities. Urban drainage failures and poorly managed waste disposal in Southwest Nigeria contribute to periodic outbreaks, particularly in high-density, low-income settlements.

Findings from this review reaffirm the crucial role of WASH (Water, Sanitation, and Hygiene) programs in mitigating cholera transmission. Government-led and NGO-backed initiatives, such as the Key Household Hygiene Practices (KHHP) and various WASH programs, have played a significant role in increasing hygiene awareness and reducing open defecation rates. However, the study highlights gaps in implementation, funding limitations, and inconsistent policy execution, particularly in rural and underserved areas. Some regions continue to struggle with open defecation despite KHHP initiatives, due to lack of community buy-in and inadequate enforcement of sanitation laws. Inconsistent clean water supply in peri-urban and rural areas reduces the long-term impact of WASH programs. Political instability and bureaucratic inefficiencies hinder the sustained implementation of sanitation projects, especially in conflict-affected regions. Without strengthening and scaling WASH infrastructure, Nigeria will continue to experience recurrent cholera epidemics, particularly in areas with poor environmental management (Perez-Saez *et al.,* 2022).

Socioeconomic disparities significantly compound the risk of cholera transmission, particularly in low-income urban slums and conflict-affected regions. The research findings indicate that Internally Displaced Persons (IDP) camps in Northeast Nigeria are among the most vulnerable environments for cholera outbreaks, primarily due to overcrowding and poor sanitation, where limited resources make it difficult to implement hygiene practices. A lack of access to clean water supplies forces residents to rely on contaminated surface water for drinking and domestic use. Inadequate humanitarian support, exacerbated by conflict-related logistical challenges, leaves IDP populations without proper medical care and preventive measures (Charnley *et al.,* 2022).

Similarly, the study identifies high cholera prevalence in urban slums across Southwest Nigeria, particularly in Lagos and Ibadan, where poor sanitation and overreliance on informal water vending systems contribute to persistent outbreaks. Findings suggest that unregulated water vending is a major public health risk, particularly in Southeast Nigeria, where residents in cities like Onitsha and Aba depend heavily on untreated water sources for daily consumption. Informal water markets remain unregulated, exposing consumers to contaminated water supplies. Poverty limits access to safer water alternatives, forcing low-income populations to prioritize affordability over safety. Low literacy rates and inadequate health education in these areas hinder the adoption of safe water handling practices. The study underscores the need for stronger urban sanitation policies, better-regulated water distribution, and targeted public health education campaigns to curb cholera transmission in these regions (Baličević *et al.,* 2023).

Behavioral factors continue to play a major role in sustaining cholera outbreaks. The study confirms that street-vended food, traditional water storage methods, and poor hand hygiene contribute significantly to cholera transmission in urban and rural settings. Consumption of unhygienically prepared street food in informal markets is one of the leading causes of foodborne cholera infections in urban centers across Southwest and North Central Nigeria (Ihua *et al.,* 2024). The use of uncovered water storage containers in rural communities increases bacterial contamination risks, particularly in Northwest and Northeast Nigeria. Hand hygiene remains inconsistent in many households, with limited access to soap and clean water in resource-poor areas.

The study suggests that community-level health education programs need to be expanded and reinforced to improve behavioral practices. Programs under KHHP should be integrated into school curricula, religious institutions, and grassroots initiatives to drive lasting behavioral change. Health education on safe food handling should be prioritized in urban slums and markets, targeting both food vendors and consumers. Community-driven sanitation initiatives can improve hygiene compliance in rural areas, encouraging households to adopt safer water storage methods. Increased public health outreach through radio and social media campaigns can enhance cholera prevention awareness, particularly among low-literacy populations.

The study findings highlight serious inadequacies within Nigeria’s healthcare system, particularly in early outbreak detection and response coordination. Delayed cholera outbreak response, especially in Northeast and North Central regions, is a major challenge, as weak disease surveillance mechanisms slow containment efforts. Limited laboratory diagnostic capacity affects rapid confirmation of cholera cases, leading to inefficient epidemic management. Funding constraints hinder proactive intervention strategies, with emergency response efforts often being reactive rather than preventive.

Despite global efforts to expand Oral Cholera Vaccine (OCV) coverage, this study confirms that Nigeria continues to face low vaccine uptake, especially in high-risk regions. OCV distribution remains inconsistent, with shortages in conflict-affected areas such as Northeast Nigeria (Ayenigbara *et al.,* 2016). Public mistrust of vaccines and logistical challenges further limit vaccination campaign effectiveness. Healthcare workforce shortages in rural and underserved areas reduce the efficiency of cholera treatment and case management efforts (Ezeh *et al.,* 2022).

The findings of this study underscore the urgent need for improved healthcare planning, including strengthening emergency response teams to ensure faster outbreak containment. Enhancing OCV distribution in high-risk areas through better supply chain management and community engagement is necessary. Expanding training programs for healthcare workers, particularly in rural and conflict-affected regions, can improve cholera case management and treatment access. Integrating cholera control into Nigeria’s national health policy ensures sustained funding for WASH interventions and vaccination programs.

The study confirms that cholera transmission in Nigeria is driven by environmental, socioeconomic, behavioral, and healthcare system weaknesses. Addressing these challenges requires region-specific interventions, improved sanitation infrastructure, enhanced healthcare preparedness, and robust health education initiatives. By scaling up proven cholera prevention strategies, Nigeria can make significant progress in reducing cholera morbidity and mortality across all geopolitical zones.

#### Geopolitical Distribution of Studies

The distribution of studies across Nigeria’s six geopolitical zones highlights regional disparities in cholera research coverage.

i. Northwest and Northeast zones had the highest number of studies, reflecting frequent cholera outbreaks, weak sanitation infrastructure, and high population density in these areas. These regions experience seasonal outbreaks due to flooding, displacement from insurgencies, and poor access to healthcare.
ii. North Central had a moderate number of studies, focusing on flood-related outbreaks and agricultural water contamination.
iii. Southwest had a balanced representation, particularly in studies addressing urban slum conditions, poor waste management, and informal settlements that exacerbate cholera transmission.
iv. Southeast and South-South had fewer studies, despite documented cholera cases in coastal and riverine communities. These regions face unique challenges such as unregulated water vending, oil-related environmental degradation, and heavy reliance on surface water sources.

The disproportionate focus on northern regions indicates that research efforts have largely been driven by cholera burden, outbreak frequency, and emergency response needs. However, the relatively lower number of studies in the South-South and Southeast suggests that cholera transmission dynamics in coastal areas require further investigation.

### 4.2 Comparison with Global Literature

The findings of this systematic review align with global cholera epidemiology studies, confirming that environmental contamination, socioeconomic inequalities, and inadequate healthcare systems are universal drivers of cholera epidemics (Emmanuella *et al.,* 2021). Countries with similar climatic and socioeconomic conditions to Nigeria continue to experience recurring cholera outbreaks, often exacerbated by natural disasters, population displacement, poor sanitation infrastructure, and weak public health systems. By analyzing cholera control efforts in different parts of the world, lessons can be drawn to inform better intervention strategies in Nigeria.

In Bangladesh, seasonal monsoon flooding has been shown to significantly increase cholera transmission by contaminating water supplies and overwhelming already fragile sanitation systems. The country experiences annual floods, particularly in low-lying riverine areas, leading to the displacement of thousands of people and creating ideal conditions for cholera outbreaks (Idoga *et al.,* 2019). These conditions mirror those found in North Central Nigeria, where seasonal flooding contributes to high cholera prevalence, particularly in communities that rely on untreated surface water sources. The government of Bangladesh has implemented early warning systems and flood-resistant latrines, alongside WASH (Water, Sanitation, and Hygiene) initiatives, to mitigate the spread of cholera. The adaptation of similar interventions, such as pre-flood chlorination of water sources and improved drainage systems, could help Nigeria reduce its vulnerability to flood-induced cholera outbreaks.

The role of socioeconomic disparities in cholera transmission is evident in Haiti and India, where poverty, urban congestion, and limited access to clean water continue to drive outbreaks (Tarh, 2020). In Haiti, particularly in Port-au-Prince, rapid urbanization has led to overcrowded informal settlements where sanitation infrastructure is practically nonexistent. These conditions are similar to those in urban slums in Southwest Nigeria, such as Lagos and Ibadan, where cholera remains endemic due to inadequate waste disposal and reliance on contaminated water supplies (Faruque *et al.,* 2019). In both cases, informal housing developments have outpaced the ability of municipal authorities to provide clean water and sanitation facilities. The Haitian government, in collaboration with international organizations, has undertaken large-scale cholera vaccination campaigns and constructed decentralized waste treatment sites. Nigeria could benefit from adopting similar approaches by prioritizing community-led sanitation projects in urban slums and expanding cholera vaccination programs in vulnerable communities.

Research from Latin America and Southeast Asia highlights the role of cultural and behavioral practices in cholera transmission. In many countries within these regions, the widespread consumption of street food and the reliance on informal water vending heighten exposure risks, particularly in low-income areas with poor regulatory oversight (Ngogo *et al.,* 2023). A similar trend is observed in Nigeria, particularly in the Southeast, where informal water vending remains the primary drinking water source for a significant portion of the population. The lack of quality control in this sector results in a high risk of cholera contamination. Efforts in Latin American countries such as Peru and Brazil have focused on public health education campaigns targeting street vendors and informal water suppliers, as well as the implementation of stricter sanitation regulations. Nigeria could enhance its cholera control efforts by improving regulatory frameworks for informal water vendors and launching public awareness campaigns emphasizing safe food and water handling practices.

Unlike Nigeria, some countries have successfully established robust cholera prevention and response mechanisms, significantly reducing mortality rates associated with outbreaks. Vietnam and South Korea have implemented early-warning surveillance systems that enable the rapid detection and containment of cholera outbreaks before they escalate (OKeeffe *et al.,* 2024). These systems integrate real-time data from hospitals, water quality monitoring stations, and meteorological services to predict and prevent outbreaks. Nigeria’s current disease surveillance system remains fragmented and reactive, often responding to outbreaks only after they have spread widely. A more proactive approach, modeled after Vietnam’s early warning system, could help Nigeria minimize cholera-related fatalities. Additionally, vaccine distribution in South Korea is streamlined through advanced logistics planning and mass immunization drives, whereas Nigeria continues to face challenges in delivering oral cholera vaccines (Ezeneme *et al.,* 2023). Implementing more efficient vaccine distribution strategies, coupled with community engagement to improve acceptance rates, could enhance Nigeria’s cholera control efforts.

The findings suggest that Nigeria could benefit significantly from adopting integrated cholera control models used in Bangladesh and Vietnam. A combination of improved WASH infrastructure, strong community engagement, and proactive vaccination programs has led to substantial reductions in cholera outbreaks in these countries (Dan *et al.,* 2019). Lessons from these regions highlight the importance of a multifaceted approach that not only addresses environmental and behavioral risk factors but also strengthens healthcare preparedness and government response mechanisms. To effectively control cholera, Nigeria must develop a sustainable strategy that incorporates best practices from around the world while tailoring interventions to its unique socioeconomic and environmental context.

Given the similarities between Nigeria and other cholera-endemic regions, policymakers should prioritize investments in sanitation infrastructure, enhance public health education, and adopt advanced disease surveillance systems to preemptively contain outbreaks. The success of countries such as Vietnam and Haiti demonstrate that with coordinated government efforts, community involvement, and international support, cholera can be controlled and ultimately eliminated. Implementing these global best practices in Nigeria would significantly contribute to reducing cholera-related morbidity and mortality, improving overall public health outcomes across the country.

### 4.3 Strengths and Limitations of the Review

#### Strengths

i. **Comprehensive Regional Analysis**: This review systematically examined cholera transmission across all six geopolitical zones in Nigeria, providing a broad yet detailed analysis of region-specific risk factors.
ii. **Multifactorial Approach**: Unlike previous studies that focused on individual risk factors, this review considers environmental, socioeconomic, cultural, and healthcare-related determinants, offering a holistic perspective on cholera transmission.
iii. **Alignment with Global Research**: The findings were compared with international cholera research, ensuring that conclusions were contextualized within the broader epidemiological landscape.
iv. **Policy Relevance**: By identifying key healthcare system challenges, this review highlights specific areas for policy intervention, particularly in outbreak response and vaccine accessibility.

#### Limitations

i. **Limited Number of Included Studies**: Due to strict eligibility criteria, only 40 studies were included in this review, which may not fully capture all regional variations in cholera transmission.
ii. **Potential Publication Bias**: The review primarily relied on peer-reviewed and publicly available studies, which may exclude unpublished government reports and field data, potentially limiting comprehensiveness.
iii. **Heterogeneity in Study Designs**: Variability in study methodologies, sample sizes, and data collection approaches may affect the comparability of findings.
iv. **Limited Discussion on Climate Change**: Although seasonal flooding was identified as a risk factor, this review does not extensively assess the long-term effects of climate change on cholera outbreaks, which warrants further investigation.
v. **Challenges in Policy Implementation**: While this review highlights deficiencies in Nigeria’s cholera control policies, it does not comprehensively analyze barriers to policy enforcement, such as political constraints and funding shortages.

## 5.0 Conclusion

This systematic review has comprehensively analyzed the key factors influencing cholera transmission across Nigeria’s geopolitical zones, emphasizing the interconnected roles of environmental, socioeconomic, cultural, and healthcare system-related elements. The findings highlight that contaminated water sources, poor sanitation, population displacement, and inadequate healthcare infrastructure are major drivers of recurrent cholera outbreaks. The study underscores that while all six geopolitical zones experience cholera to varying degrees, certain regions, such as the Northwest and Northeast, are disproportionately affected due to weak sanitation systems, open defecation, and unreliable access to potable water. The South-South region faces unique environmental challenges, particularly oil-related water pollution, while urban slums in the Southwest and Southeast remain vulnerable due to unregulated water vending and poor waste disposal. These findings reaffirm the urgent need for targeted cholera mitigation strategies tailored to regional-specific risks.

The study also underscores the significant impact of socioeconomic disparities on cholera vulnerability, particularly in low-income urban settlements and IDP camps in the Northeast. Overcrowding, lack of access to clean drinking water, and reliance on informal water supply systems continue to fuel cholera transmission. In the Southwest and Southeast, the dominance of informal water vending and street food consumption further exacerbates cholera outbreaks, pointing to a need for stricter regulatory oversight and improved sanitation policies. Despite government and NGO-led WASH interventions, the findings indicate that gaps in funding, inconsistent implementation, and weak policy enforcement limit the effectiveness of these programs. Strengthening community-based sanitation programs, integrating cholera prevention education into school curricula, and providing sustained investment in clean water infrastructure are crucial to addressing these socioeconomic disparities.

Global comparisons reveal that Nigeria’s cholera burden is not unique, as countries such as Bangladesh, Haiti, and India face similar environmental and socioeconomic risk factors. However, the review indicates that Nigeria lags behind in cholera surveillance, rapid outbreak response, and vaccine coverage. Countries like Vietnam and South Korea have successfully implemented early-warning disease surveillance systems and mass vaccination campaigns, significantly reducing cholera mortality rates. The study suggests that Nigeria must adopt a more proactive approach to outbreak prevention, including enhanced vaccine distribution, improved epidemiological data collection, and faster outbreak response mechanisms. Lessons from other countries emphasize that a combination of improved WASH infrastructure, effective vaccine distribution, and public health engagement is key to achieving long-term cholera control.

Addressing cholera in Nigeria requires a multifaceted, region-specific strategy that integrates public health policy reforms, infrastructure development, and behavioral change initiatives. The study’s findings reinforce the urgent need for sustained investment in WASH programs, expanded oral cholera vaccination efforts, and stronger government coordination in epidemic preparedness and response. By scaling up intervention programs, strengthening regulatory frameworks, and fostering community participation, Nigeria can significantly reduce cholera transmission, improve public health resilience, and work towards the eventual elimination of cholera outbreaks.

## 6.0 Recommendation

### 6.1 Public Health Recommendations for Future Cholera Control

Despite notable progress, challenges remain in achieving universal access to cholera prevention and treatment measures. The following strategies are recommended to enhance cholera control efforts:

i. Expanding OCV Coverage in High-Risk Areas: Increased investment in oral cholera vaccine distribution in conflict-affected and hard-to-reach areas.
ii. Strengthening Disease Surveillance and Early Detection: Enhancing real-time data tracking and response systems to contain outbreaks more effectively.
iii. Scaling Up WASH Infrastructure: Prioritizing sanitation and clean water projects, especially in IDP camps and informal settlements.

Regional Policy Implementation: Adopting geopolitical-specific cholera control strategies, ensuring that interventions address the unique challenges of each region.

### 6.2 Public Health Implications and Recommendations

Rectifying WASH inadequacies is essential for cholera prevention and management in Nigeria. The results highlight the pressing necessity for cohesive intervention measures, encompassing:

i. Investment in Water Infrastructure: Enhancing borehole initiatives, upgrading urban water treatment facilities, and guaranteeing the provision of safe drinking water in high-risk regions.
ii. Sanitation Improvement Programs: Executing community-driven sanitation efforts to mitigate open defecation and improve waste disposal systems.
iii. Hygiene Promotion and Public Awareness: Implementing extensive educational efforts to promote handwashing, proper food handling, and water purification methods in communities at high risk for cholera.
iv. Government and Policy Intervention: Enhancing cholera surveillance systems and implementing policy-driven strategies to improve sanitation and hygiene, particularly in internally displaced persons (IDP) camps and informal settlements.

### 6.3 Public Health Recommendations

Addressing gaps in healthcare preparedness requires a multifaceted approach:

i. Strengthening Healthcare Infrastructure: Increasing the number of cholera treatment centers in high-risk areas, particularly in conflict-affected regions.
ii. Reducing Response Time: Enhancing early detection mechanisms, improving emergency response logistics, and establishing regional outbreak control teams.
iii. Expanding Vaccine Distribution: Implementing targeted vaccination campaigns, ensuring consistent supply chains, and addressing vaccine hesitancy in vulnerable communities.
iv. Improving Coordination Between Government and NGOs: Strengthening partnerships to facilitate better healthcare access in IDP camps and rural communities.

## Data Availability

All data produced in the present work are contained in the manuscript

## Notes

### Competing Interest Statement

The authors have declared no competing interest.

### Funding Statement

This study did not receive any funding

